# Associations between circulating metabolites and incidence of breast, prostate, lung and colorectal cancers in the HUNT Study and UK Biobank

**DOI:** 10.1101/2025.10.22.25338179

**Authors:** Marion Denos, Abhibhav Sharma, Bjørn Olav Åsvold, Alya G.A. Arham, George Davey Smith, Nicholas J. Timpson, Mari Løset, Ben Michael Brumpton, Julia Debik, Guro F. Giskeødegård

## Abstract

**Background:** Breast, prostate, lung and colorectal cancers are the most common cancers in both women and men globally. The role of circulating metabolites, including lipoprotein subfractions, in the development of these cancers remains unclear.

**Objective:** This prospective cohort study, including two large population-based cohorts, aimed to explore the associations between circulating metabolites and the incidence of breast, prostate, lung and colorectal cancers.

**Methods:** From the Trøndelag Health Study (HUNT) and the UK Biobank (UKBB), 132 lipoprotein subfractions and metabolites, quantified using nuclear magnetic resonance spectroscopy, were available for participants (n=16,877 in HUNT3, n=260,805 in UKBB). Cox regression was used to investigate the associations with incident cancers.

**Results:** During a mean follow-up time of 14 years, 549 women and 741 men developed breast (n= 283), prostate (n=452), lung (n=175) or colorectal (n=380) cancer in HUNT3, while 6679 women and 8248 men developed breast (n=4516), prostate (n=5576), lung (n=1985) or colorectal (n=2850) cancer in UKBB. In the UKBB, higher levels of several subfractions of lipoproteins including VLDL and LDL were associated with a slight decreased incidence of prostate and lung cancers (cholesterol in large LDL: HR per 1-SD increment 0.90, 95% CI 0.86-0.94 for lung cancer). Similar associations were observed between albumin, some fatty acids, and lung cancer. However, no clear associations were observed in HUNT3.

**Conclusion:** Several lipoprotein subfractions and circulating metabolites were associated with a reduced incidence of lung cancer, predominantly in UKBB. Although no conclusive evidence was found for a specific lipid or metabolite profile associated with the incidence of the most common cancers, the observed associations were generally consistent across cohorts. Our results highlight promising directions for future research into metabolic biomarkers and their potential role in cancer prevention.

## Introduction

Globally around 20 million new cancer cases were reported in 2022, and predictions estimate this number to rise to 35 million by 2050 (1). Breast cancer (BC) is the most common cancer in women, while prostate cancer (PCa) ranks second in men (1). Lung cancer (LC) and colorectal cancers (CRC) are also among the most frequently diagnosed cancers in both sexes(1). Although several modifiable risk factors have been associated with cancer incidence, they only partially account for the observed rates. In BC, factors related to reproductive history and lifestyle have been identified, yet they only explain a small proportion of cases in high-income countries (2–7). For PCa, few lifestyle and environmental factors have been established; age, ethnicity, family history and genetic susceptibility are recognized contributors, while associations with smoking and body weight remain inconclusive (1, 3, 8–10). LC is primarily attributed to tobacco smoking, and is often diagnosed at an advanced stage, resulting in poor survival rates. This underscores the need for biomarkers that enable earlier detection (1, 11). CRC risk is associated with factors such as age, smoking, alcohol consumption and diet (12). However, its typically slow progression and lack of early symptoms pose challenges for timely diagnosis. While prostate-specific antigen is used as a diagnostic tool for PCa, it does not predict cancer risk. To date, no clinically established circulating risk markers exist for these four major cancer types.

Metabolomics captures the dynamic interaction between an individual and the surrounding environment, revealing insights closely related to the phenotype (13, 14). The metabolite profile thus reflects the biological condition, providing a snapshot of the organism’s current state. Metabolites found in human biofluids may serve as biomarkers of current health and future disease risk, and previous studies have shown the potential of metabolomic biomarkers for disease risk (15–17). Altered cellular metabolism is a well-established hallmark of cancer (18). In particular, changes in the levels of specific lipids and small-molecular have been shown to support anaerobic glycolysis, thereby meeting the elevated energy and biosynthetic demands of tumor growth (19, 20). However, the role of circulating metabolites and lipids in cancer risk remains inconclusive. Epidemiological studies have found a strong link between elevated blood lipid levels and a higher incidence and recurrence of PCa, while inhibiting lipid biosynthesis with statins has been shown to reduce mortality of BC, LC and colon cancer (21–23). Mass spectrometry (MS) and nuclear magnetic resonance (NMR) based case-control studies have reported varying results on associations between metabolic profiles of prediagnostic blood samples and incidence of BC, PCa, LC and CRC (24–37). Our previous case-control study, based on the second Trøndelag Health Study (HUNT2), found that high levels of very low-density lipoprotein (VLDL) subfractions in blood were associated with a reduced risk of BC in premenopausal women and revealed alterations in lipid metabolism many years before BC diagnosis (29). Recently, studies by Buergel et al. and Julkunen et al. utilized the initial release of UK Biobank metabolomics data from ∼118,000 participants (38, 39). They reported no or weak associations between free circulating metabolite levels and incidence of BC, PCa or CRC while they identified several biomarkers for LC incidence (38, 39). Both studies did not account for reproductive factors as confounders in women, which might influence the associations. In contrast, other case-control studies with a prospective design have identified metabolite risk factors for PCa (32, 40, 41). Thus, studies exploring the association between metabolic biomarkers and risk of BC, PCa, LC and CRC have shown inconsistent results and require further investigation.

In this prospective cohort study, we aimed to investigate the associations between circulating lipoprotein subfractions and small-molecular metabolites and incidence of BC, PCa, LC and CRC using data from two large population-based cohorts for independent validation: The Trøndelag Health Study (HUNT) in Norway and the UK Biobank including Phase 1 and Phase 2 metabolomic data releases.

## Methods

### Study populations

#### HUNT3

The Trøndelag Health Study (HUNT) is a longitudinal population-based cohort study in Norway, including data from questionnaires, clinical measurements, and biological materials (42). The study enrolled participants aged ≥20 years in 4 health surveys: HUNT1 (1984–1986), HUNT2 (1995–1997), HUNT3 (2006–2008), and HUNT4 (2017–2019) (43). Of the 50,800 participants in HUNT3 (participation rate of 54.1%), a subset of 17,210 participants had serum metabolomics data quantified by NMR spectroscopy, generated by Nightingale Health (44). Each participant was followed up from the date of participation in HUNT3 until the date of first diagnosis of BC, PCa, LC or CRC, the date of death or emigration from Norway, or the end of the follow-up period in December 2023, whichever came first. Diagnoses of BC, PCa, LC and CRC were obtained from the Cancer Registry of Norway. Information on death was collected from the Norwegian Cause of Death Registry. Among the 17,210 participants, we excluded those with a prior diagnosis of the cancer under study (breast, prostate, lung, or colorectal) before inclusion into HUNT3, leaving on average 16,877 participants in the study population.

#### UK Biobank (UKBB)

UKBB is a large population-based prospective study of adults aged 40-69 years recruited in 2006-2010 (45). Among the 9.2 million individuals invited to the study, around 500,000 (5.5%) participated. A large range of health data, including questionnaires and blood samples, was collected in 22 assessment centers throughout the United Kingdom (UK) (45). Metabolic profiling of all participants’ plasma samples is underway using the Nightingale Health NMR platform, and here we used the second release of metabolomics data of 274,352 participants (46). Participants were followed up from the participation date in UKBB to the date of first diagnosis of BC, PCa, LC or CRC, the date of death or lost to follow-up, or the end of the follow-up period in June 2022, whichever came first. UKBB linked data from the National Cancer and Death Registries to its database. Among the 268,047 participants, we excluded those with a prior diagnosis of the cancer under study (breast, prostate, lung, or colorectal) before inclusion into UKBB, leaving on average 260,805 participants in the study population.

### Metabolomics data

#### HUNT3

Metabolomics analysis of serum samples from 17,210 participants in HUNT3 was generated by NMR spectroscopy using the Nightingale Health NMR biomarker platform (Supplementary Table S1). The serum samples were non-fasting, with an average of 3 hours since the last meal. The samples were analyzed in two batches. Batch 1 (n = 6201) was analyzed using a 600 MHz Bruker AVANCE spectrometer, while batch 2 (n = 11009) was analyzed using one 500 MHz spectrometer and one 600 MHz spectrometer. Batch 2 was adjusted for instrument effects using repeated samples from 96 individuals that were measured at both instruments, by multiplying samples analyzed on 500 MHz by a correction factor per variable, resulting in equal median values for the repeated samples. Finally, batch 2 was adjusted towards batch 1 by multiplying with a correction factor per variable that resulted in equal means in the two batches. This study includes 132 metabolic variables quantified in absolute levels. The variables indicating ratios, percentages and size based on diameters were excluded from the analysis. The metabolic variables include lipoprotein subclasses, fatty acids, and small-molecular metabolites, such as amino acids, ketone bodies, glycolysis metabolites, aromatic and branched chain amino acids. Lipid concentrations and composition were measured for 14 lipoprotein subclasses, including triglycerides, phospholipids, total cholesterol, cholesteryl esters, and free cholesterol, as well as total lipid concentration for each subclass. The average biomarker detection rate was >99% in the serum samples.

#### UKBB

Nightingale Health has performed metabolic profiling of plasma samples for all UKBB participants. The plasma samples were non-fasting, with an average of 4 hours since the last meal. Measurements took place in Finland, using 500 MHz NMR Bruker AVANCE spectrometers in three different phases, as described elsewhere (39, 46). The first data release covered metabolic variables from a random selection of 118,000, the second release, which we used in the current study, added 157,000 samples, and the remaining were analyzed in the third phase, which is not yet publicly available. Any observations with more than 10% missing metabolite values or flagged with “Low protein” (indicating more severe sample dilution) were removed before imputation. We conducted kNN imputation for data missing at random, and quantile regression imputation of left-censored data (QRILC) for data missing not at random. The same metabolic variables as described for HUNT3 were quantified in UKBB, of which 132 were included in this study.

### Covariates

Weight and height were measured by health professionals during the clinical examination. Body mass index (BMI) was calculated as weight divided by the squared value of height (kg/m²) and analyzed as a continuous variable. Information on sociodemographic (age, sex, marital status), lifestyle (smoking status, alcohol consumption) and reproductive factors (age at menarche, age at menopause, number of live births) was collected in questionnaires or through interviews in both cohorts. Covariates were categorized as follows: smoking status (never, former, current), alcohol consumption (never, 1-3, ≥4 times/month), and marital status (married, unmarried).

Women with unknown menopausal status were defined as premenopausal if they were below 51 years old at recruitment and as postmenopausal if aged 51 or above. The cut-off value of 51 years was chosen based on a large study of over 300,000 Norwegian women (47). The variable IDs from both biobanks are provided in Supplementary Table S2.

### Cancer ascertainment

Participants in HUNT3 was linked to the Cancer Registry of Norway using the 11-digit personal identification number. The Cancer Registry of Norway provides accurate, close-to-complete and timely data (48). In UKBB, participants were linked to the national cancer registry which consolidates information from regional cancer centres and whose data are considered the gold standard for ascertaining cancer outcomes in the UK (49, 50). Cancer diagnoses were classified according to the Tenth Revision of the International Classification of Diseases (ICD10) as follows: BC (C50), PCa (C61), LC (C33-C34) and CRC (C18-C20) (51).

### Statistical analysis

Baseline characteristics of women and men in each cohort were described using mean and standard deviation (SD) for continuous variables, and number of participants (%) for categorical variables. Metabolomics data from UKBB were corrected for the NMR spectrometer used for the measurements, following the procedure by Julkunen et al. (39). We fit a linear regression model with log-transformed concentration as the outcome and spectrometer as the predictor. Scaled residuals from this regression were used as predictors in the analyses. For each metabolite, concentrations outside four interquartile ranges from the median were considered outliers and excluded from the analyses.

Cox proportional hazard (PH) models were used to explore the potential associations of metabolomics data with incidence of BC (in women), PCa (in men), LC and CRC, separately. Analyses were sex specific for BC and PCa. Crude and adjusted hazard ratios (HR) with 95% confidence intervals (CIs) were calculated with age as the time scale. The HRs were reported per SD increment in the log-transformed metabolite concentrations. The models were adjusted for potential confounders, including BMI, assessment center (for UKBB), smoking status, and alcohol consumption. Additionally, analyses for BC were adjusted for age at menarche and number of live births, while analyses for PCa were adjusted for marital status, and analyses for LC and CRC were adjusted for sex. We also conducted stratified analyses based on women’s menopausal status for BC. P-values were corrected for multiple testing using the Benjamini-Hochberg approach. To assess the robustness of the findings, we performed sensitivity analyses: (1) we restricted the age at participation in HUNT3 from 40 to 69 years old to match the UKBB age frame; (2) we excluded all participants with any type of cancer before baseline.

Partial least squares discriminant analysis (PLS-DA) was utilized to evaluate the comparability of NMR data between the UKBB and HUNT3. PLS-DA was applied to metabolomic data from each cohort to discriminate premenopausal and postmenopausal women. Five-fold cross-validation was applied for model training. By examining the loadings of the most explanatory latent variable, the study assessed the consistency of metabolic differences across the two population cohorts. All statistical analyses were performed with R statistical software version 4.2.1 (R Foundation for Statistical Computing, Vienna, Austria). Cox-PH analysis was performed using the *survival* package in R (52). PLS-DA was performed using the *mixOmix* package in R (53).

## Results

### Population characteristics

Table 1 outlines the characteristics of participants in the two cohorts separated by sex. The mean age of participants was around 52 years in HUNT3 and 56 years in UKBB. HUNT3 had a higher number of smokers, while UKBB participants had on average a more frequent alcohol consumption. Moreover, women in HUNT3 had an earlier age of menopause and a higher number of live births.

**Table 1.** Baseline characteristics of participants in HUNT3 and UK Biobank.

|  | HUNT3 |  | UK Biobank |  |
| --- | --- | --- | --- | --- |
|  | Women | Men | Women | Men |
| Number of participants | 9129 | 7748 | 139,684 | 121,121 |
| Female breast cancer cases | 283 (3.1) |  | 4516 (3.2) |  |
| Prostate cancer cases |  | 452 (5.8) |  | 5576 (4.6) |
| Lung cancer cases | 91 (1.0) | 84 (1.1) | 950 (0.7) | 1035 (0.8) |
| Colorectal cancer cases | 175 (1.9) | 205 (2.6) | 1213 (0.8) | 1637 (1.3) |
| Age at cancer diagnosis (yrs) | 68.2 ± 12.2 | 71.8 ± 8.9 | 64.6 ± 8.2 | 67.9 ± 6.4 |
| Age at participation (yrs) | 51.8 ± 16.0 | 52.4 ± 15.4 | 56.3 ± 8.0 | 56.6 ± 8.2 |
| Fasting time at blood collection (hours) | 2.7 ± 2.1 | 2.9 ± 2.1 | 3.7 ± 2.3 | 3.9 ± 2.6 |
| Body mass index (kg/m<sup>2</sup>) | 26.8 ± 4.8 | 27.4 ± 3.7 | 27.1 ± 5.2 | 27.8 ± 4.2 |
| Smoking status |  |  |  |  |
| never | 4006 (43.9) | 3195 (41.2) | 83,066 (59.5) | 59,072 (48.8) |
| former | 2566 (28.1) | 2617 (33.8) | 43,484 (31.1) | 46,429 (38.3) |
| current | 2293 (25.1) | 1769 (22.8) | 12,469 (8.9) | 15,021 (12.4) |
| missing | 264 (2.9) | 167 (2.2) | 545 (0.4) | 489 (0.5) |
| Alcohol consumption (times/month) |  |  |  |  |
| never | 478 (5.2) | 190 (2.5) | 12,940 (9.3) | 7605 (6.3) |
| 1 - 3 | 5774 (63.2) | 4027 (52.0) | 39,050 (28.0) | 19,556 (16.2) |
| ≥4 | 2600 (28.5) | 3413 (44.1) | 87,439 (62.6) | 93,692 (77.4) |
| missing | 277 (3.0) | 118 (1.5) | 134 (0.1) | 157 (0.1) |
| Age at menarche (yrs) | 13.2 ± 1.4 |  | 12.5 ± 2.9 |  |
| Age at menopause (yrs) | 48.6 ± 5.8 |  | 46.4 ± 13.4 |  |
| Number of live births (n) | 2.5 ± 1.1 |  | 1.8 ± 1.2 |  |
| Marital status |  |  |  |  |
| married | 5233 (57.3) | 4776 (61.7) | 98,522 (70.5) | 93,912 (77.5) |
| unmarried | 3882 (42.5) | 2956 (38.1) | 41,162 (29.4) | 27,209 (22.4) |

Data are given as mean ± standard deviation or as number of individuals (%)

Among 16,877 participants in HUNT3, 283 women developed BC, 452 men developed PCa, 175 participants developed LC, and 380 participants developed CRC, during a follow-up time of 14 years. Among 260,805 participants in the UKBB, 4516 women developed BC, 5576 men developed PCa, 1985 participants developed LC, and 2850 participants developed CRC, over a 14-year follow-up period.

### Lipoprotein and small molecule metabolite associations with cancer incidence

Hazard ratio plots from fully adjusted Cox regression models show the associations between metabolic variables and the incidences of BC (Figure 1), PCa (Figure 2), LC (Figure 3) and CRC (Figure 4).

**Figure 1.**
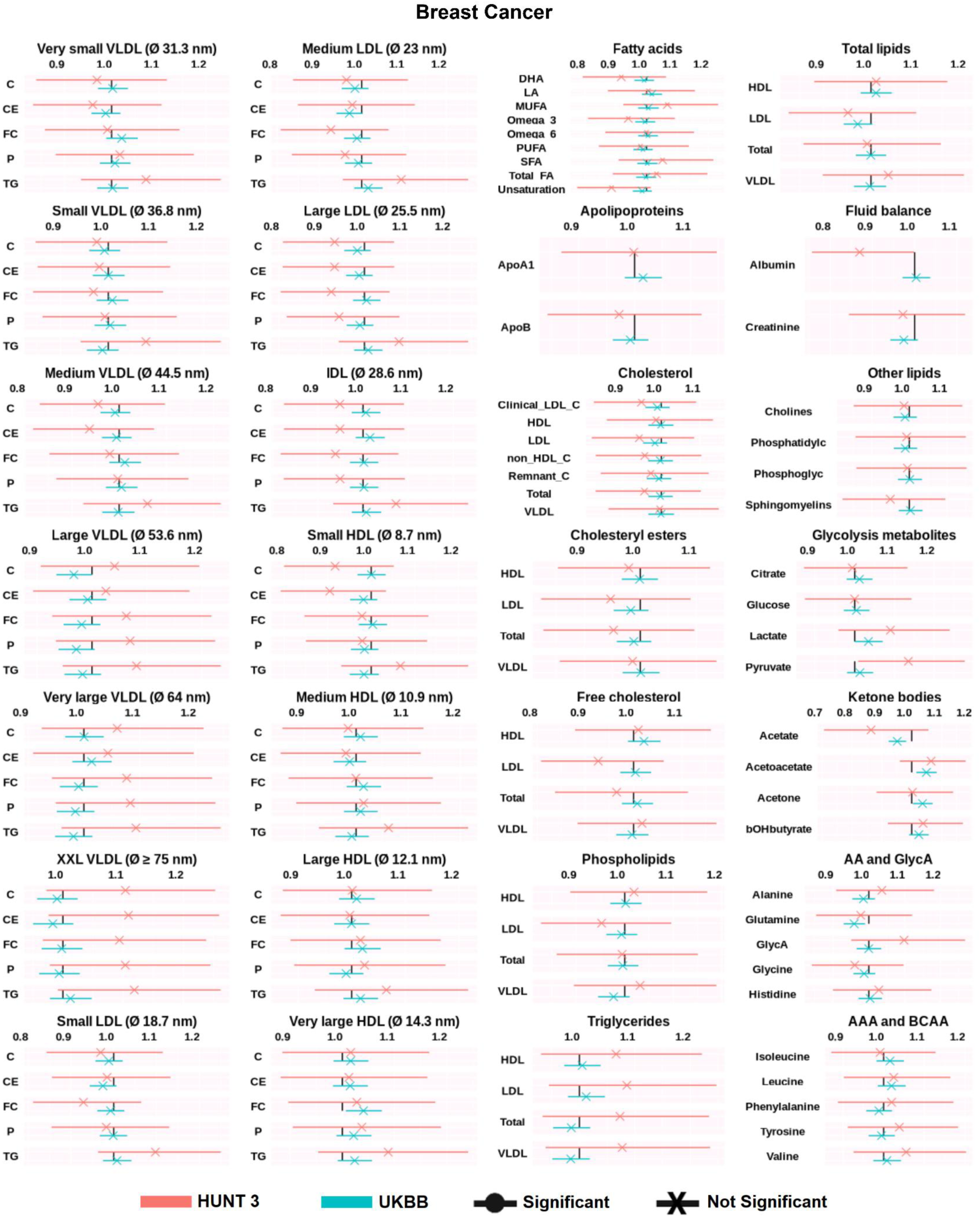
Hazard ratios (HR) of circulating metabolites in relation to breast cancer incidence were assessed in HUNT3 (events = 283, n = 9129) and the UKBB (events = 4516, n = 139,684). The data are presented as HRs per 1-SD increment with 95% confidence intervals (CIs), based on SD-scaled concentrations. Cox proportional hazards regression models were adjusted for UKBB assessment center (for UKBB), BMI, alcohol consumption, smoking status, menarche status, and number of live births, with age used as the timescale. AA=Amino acids, AAA = Aromatic amino acids, ApoA1 = Apolipoprotein A1, ApoB = Apolipoprotein B, bOHbutyrate = Beta-hydroxybutyrate, BCAA = Branched-chain amino acids, CE = Cholesteryl esters, C = Cholesterol, DHA = Docosahexaenoic acid, FC = Free cholesterol, GlycA = Glycoprotein Acetylation, HDL = High-density lipoprotein, LA = Linoleic acid, LDL = Low-density lipoprotein, MUFA = Monounsaturated fatty acids, Phosphatidylc = Phosphatidylcholine, Phosphoglyc = Phosphoglycerides, P = Phospholipids, PUFA = Polyunsaturated fatty acids, SFA = Saturated fatty acids, TG = Triglycerides, Total FA = Total fatty acids, VLDL = Very low-density lipoprotein, XXL = Extremely large, Ø = Diameter.

**Figure 2.**
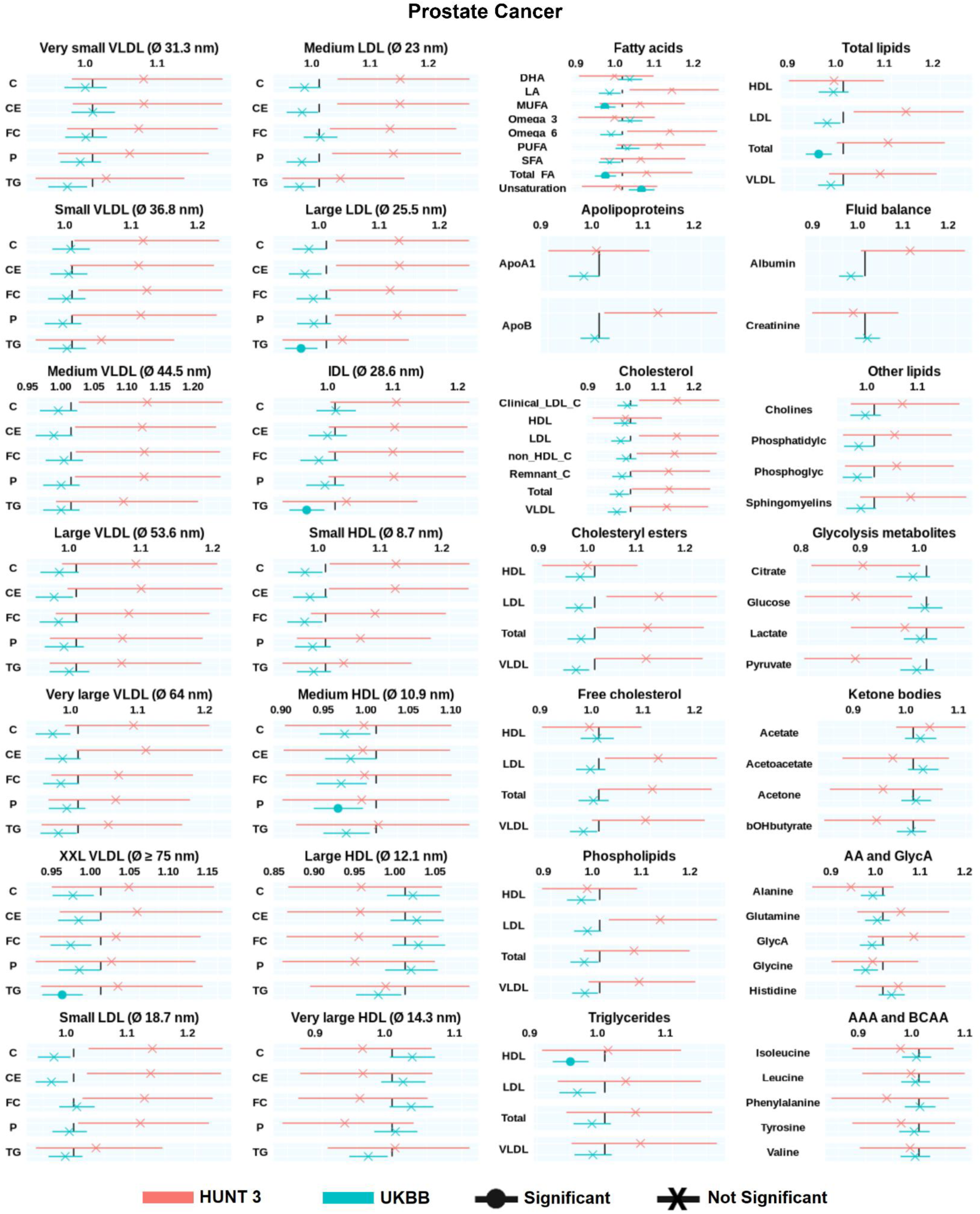
Hazard ratios (HR) of circulating metabolites in relation to prostate cancer incidence were assessed in HUNT3 (events = 452, n = 7,748) and the UKBB (events = 5,576, n = 121,121). The data are presented as HRs per 1-SD increment with 95% confidence intervals (CIs), based on SD-scaled concentrations. Cox proportional hazards regression models were adjusted for UKBB assessment center (for UKBB), BMI, alcohol consumption, smoking status, marital status, with age used as the timescale. AA=Amino acids, AAA = Aromatic amino acids, ApoA1 = Apolipoprotein A1, ApoB = Apolipoprotein B, bOHbutyrate = Beta-hydroxybutyrate, BCAA = Branched-chain amino acids, CE = Cholesteryl esters, C = Cholesterol, Clinical_LDL_C, DHA = Docosahexaenoic acid, FC = Free cholesterol, GlycA = Glycoprotein Acetylation, HDL = High-density lipoprotein, LA = Linoleic acid, LDL = Low-density lipoprotein, MUFA = Monounsaturated fatty acids, Phosphatidylc = Phosphatidylcholine, Phosphoglyc = Phosphoglycerides, P = Phospholipids, PUFA = Polyunsaturated fatty acids, SFA = Saturated fatty acids, TG = Triglycerides, Total FA = Total fatty acids, VLDL = Very low-density lipoprotein, XXL = Extremely large, Ø = Diameter.

**Figure 3.**
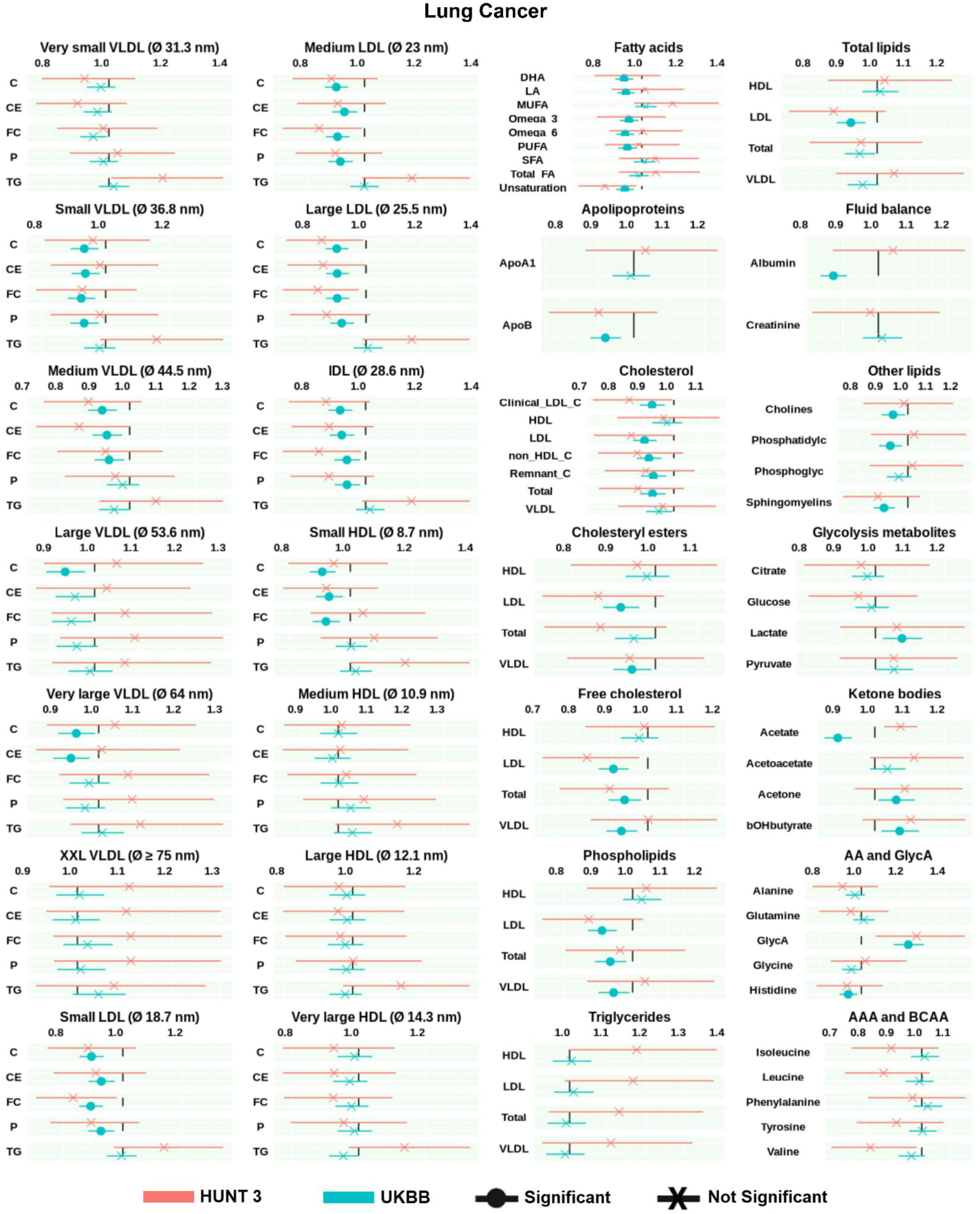
Hazard ratios (HR) of circulating metabolites in relation to lung cancer incidence were assessed in HUNT3 (events = 175, n = 16,877) and the UKBB (events =1985, n = 260,805). The data are presented as HRs per 1-SD increment with 95% confidence intervals (CIs), based on SD-scaled concentrations. Cox proportional hazards regression models were adjusted for sex, UKBB assessment center (for UKBB), BMI, alcohol consumption, smoking status, with age used as the timescale. AA=Amino acids, AAA = Aromatic amino acids, ApoA1 = Apolipoprotein A1, ApoB = Apolipoprotein B, bOHbutyrate = Beta-hydroxybutyrate, BCAA = Branched-chain amino acids, CE = Cholesteryl esters, C = Cholesterol, Clinical_LDL_C, DHA = Docosahexaenoic acid, FC = Free cholesterol, GlycA = Glycoprotein Acetylation, HDL = High-density lipoprotein, LA = Linoleic acid, LDL = Low-density lipoprotein, MUFA = Monounsaturated fatty acids, Phosphatidylc = Phosphatidylcholine, Phosphoglyc = Phosphoglycerides, P = Phospholipids, PUFA = Polyunsaturated fatty acids, SFA = Saturated fatty acids, TG = Triglycerides, Total FA = Total fatty acids, VLDL = Very low-density lipoprotein, XXL = Extremely large, Ø = Diameter.

**Figure 4.**
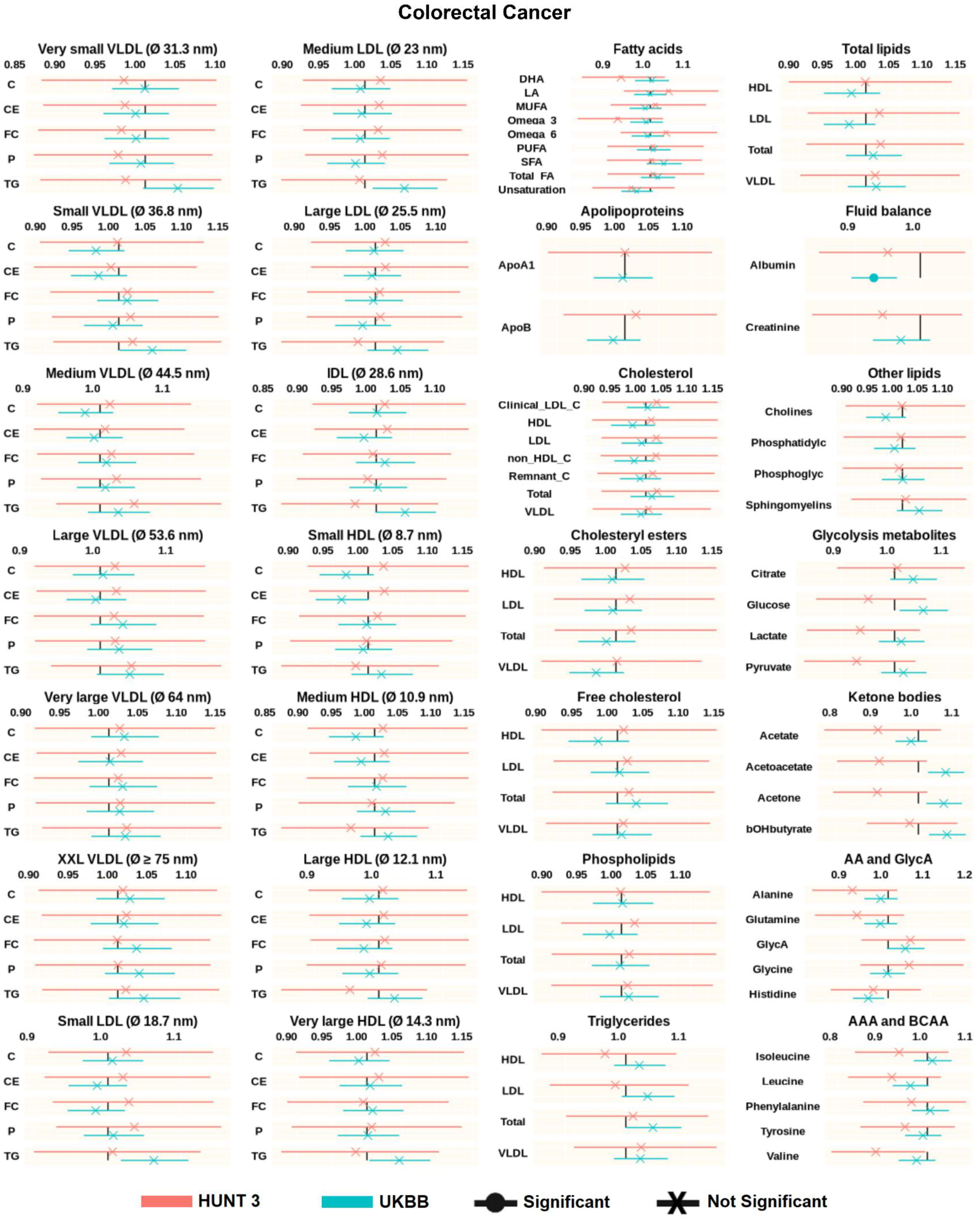
Hazard ratios (HR) of circulating metabolites in relation to colorectal cancer incidence were assessed in HUNT3 (events = 380, n = 16,877) and the UKBB (events = 2850, n = 260,805). The data are presented as HRs per 1-SD increment with 95% confidence intervals (CIs), based on SD-scaled concentrations. Cox proportional hazards regression models were adjusted for sex, UKBB assessment center (for UKBB), BMI, alcohol consumption, smoking status, with age used as the timescale. AA=Amino acids, AAA = Aromatic amino acids, ApoA1 = Apolipoprotein A1, ApoB = Apolipoprotein B, bOHbutyrate = Beta-hydroxybutyrate, BCAA = Branched-chain amino acids, CE = Cholesteryl esters, C = Cholesterol, Clinical_LDL_C, DHA = Docosahexaenoic acid, FC = Free cholesterol, GlycA = Glycoprotein Acetylation, HDL = High-density lipoprotein, LA = Linoleic acid, LDL = Low-density lipoprotein, MUFA = Monounsaturated fatty acids, Phosphatidylc = Phosphatidylcholine, Phosphoglyc = Phosphoglycerides, P = Phospholipids, PUFA = Polyunsaturated fatty acids, SFA = Saturated fatty acids, TG = Triglycerides, Total FA = Total fatty acids, VLDL = Very low-density lipoprotein, XXL = Extremely large, Ø = Diameter.

No lipoprotein subfractions or circulating metabolites were associated with BC in UKBB or HUNT3 after adjusting for multiple testing (Figure 1). Hazard ratios and corresponding 95% CIs for age-adjusted and fully adjusted Cox models for BC are provided in Supplementary Data 1-2.

Several fatty acids and lipoprotein subfractions were associated with PCa in UKBB (Figure 2). Higher levels of total triglycerides in HDL were associated with a decreased PCa incidence in the UKBB cohort (HR per 1-SD increment 0.95, 95% CI 0.92-0.98). A similar trend was observed for LDL and VLDL subfractions. Fatty acids species, including monounsaturated fatty acids (MUFA) (HR 0.96, 95% CI 0.93-0.98) and total fatty acids (HR 0.96, 95% CI 0.93-0.98), were also associated with a decreased risk of PCa, while unsaturated fatty acids were associated with an increased risk of PCa (HR = 1.05, 95% CI 1.02-1.08). There was no association between metabolic variables and PCa in HUNT3. Notably, the point estimates were in the opposite direction for some metabolic variables in HUNT3 compared to those observed in the UKBB. However, the 95% CIs were wide in HUNT3 due to its smaller sample size (Supplementary Data 1-2).

As presented in Figure 3, total cholesterol, total free cholesterol, and total phospholipids were inversely associated with LC incidence in the UKBB (total cholesterol: HR per 1-SD increment 0.93, 95% CI 0.88-0.97). These inverse associations were specifically evident in small and medium VLDL, all LDL and IDL particles. Apolipoprotein B (ApoB) was associated with a reduced LC risk (HR per 1-SD increment 0.92, 95% CI 0.88-0.96). Additionally, higher levels of fatty acids, including docosahexaenoic acid, linoleic acid, omega-3 and –6, and polyunsaturated fatty acids (PUFAs), were associated with a reduced risk of LC. Similarly, albumin level was inversely associated with LC as well as circulating levels of cellular membrane-based lipids, such as choline, phosphatidylcholine, and sphingomyelins. Conversely, glycolysis-related metabolites such as lactate and free circulating ketone bodies (acetone, beta-hydroxybutyrate) were positively associated with LC incidence, while acetate was negatively associated with LC. Notably, the inflammation marker glycoprotein acetylation (GlycA) was positively associated with LC incidence (HR per 1-SD increment 1.23, 95% CI 1.15-1.30). The results from HUNT3 were consistent with UKBB, although not statistically significant after multiple testing adjustment, likely due to the smaller sample size. Hazard ratios and CIs for age-adjusted and fully adjusted Cox models for LC are provided in Supplementary Data 1-2.

Higher level of albumin was associated with a reduced risk of CRC in the UKBB (HR per 1-SD increment 0.93, 95% CI 0.89-0.96) (Figure 4). Glycolysis-related metabolites such as glucose and free circulating ketone bodies (acetoacetate, acetone, beta-hydroxybutyrate) were positively associated with CRC incidence, although these associations were not statistically significant after multiple testing correction. The inflammation marker GlycA was positively associated with CRC incidence (HR per 1-SD increment 1.04, 95% CI 1.00-1.09), similarly to its association with LC incidence. No lipoprotein subfractions or other metabolites were associated with CRC in UKBB or HUNT3. Hazard ratios and CIs for age-adjusted and fully adjusted Cox models for CRC are provided in Supplementary Data 1-2.

### Sensitivity analysis

Given the previously reported differences in serum metabolomic profiles between premenopausal and postmenopausal women (29), which were confirmed in our study (Supplementary Figure S1), we conducted stratified analyses to investigate the associations of metabolic variables with BC incidence in pre-diagnostic blood samples of premenopausal and postmenopausal women separately. Several metabolic variables were inversely associated with BCa in premenopausal women, but not in postmenopausal women, in UKBB. These included total triglyceride levels, triglycerides and cholesterols in large VLDLs, cholesterol and cholesteryl esters in LDL subfractions, all lipids in small HDL, in addition to several fatty acid (Supplementary Figure S2, Supplementary Data 1-2). No clear associations were observed in HUNT3.

Results remained similar when the analysis was performed in the age restricted HUNT3 cohort (in both raw and adjusted models) (Supplementary Figures S3–6, Supplementary Data 1-2). Moreover, the associations remained unchanged when participants with previous history of cancer at baseline were excluded (Supplementary Data 3-4).

## Discussion

This prospective study explored the association between circulating metabolites, including lipoprotein subfractions and lipids, and the incidence of BC, PCa, LC or CRC, using data from two large cohorts: HUNT3 and the UKBB. Several metabolites were associated with cancer incidence in UKBB, particularly for LC. Higher levels of triglycerides were associated with a lower risk of PCa. In contrast, higher levels of most lipid concentrations in LDL and IDL subfractions, except triglycerides, were associated with lower risk of LC. No clear associations were observed for BC before analyses were stratified based on women’s menopausal status, where several lipoprotein subfractions showed a protective association in premenopausal women. Higher levels of albumin were inversely associated with both LC and CRC, while the inflammation marker GlycA showed a positive association with both cancers. In addition, free circulating ketone bodies (acetone, beta-hydroxybutyrate) were positively associated with incidence of LC and CRC. Except for PCa, results from HUNT3 were generally consistent with UKBB, but with wider confidence intervals, likely due to smaller sample size.

Lipids, including fats, oils, phospholipids, sterols, and triglycerides, are essential for energy storage, membrane structure, and cellular signaling (26, 28, 54–58). Lipoproteins, transporters of lipids in the blood, are classified into five types—VLDL, IDL, LDL, HDL, and chylomicrons. While LDL carries triglycerides and cholesterol from the liver to tissues in the endogenous pathway, HDL collects excess cholesterol from tissues and blood vessels and transports it back to the liver for removal (59). So far, few studies have investigated the role of lipoprotein subfractions in cancer risk. We found the level of several lipoprotein subfractions in the endogenous pathway to be inversely associated with LC. Inverse associations between LDL-cholesterol and lung cancer risk have been seen in other large prospective cohorts (60), and a Mendelian randomization study showed evidence for a causal effect of high levels of LDL-cholesterol on a reduced risk of LC (61). However, results are conflicting as a recent Mendelian randomization study reported no association between lipid subfractions and cancer risk (62). Furthermore, several prospective studies based on NMR reported inverse associations of free circulating lipids and fatty acids with long-term PCa risk (26, 55–58). In line with these studies, we observed negative associations between the levels of total lipids or total fatty acids and PCa risk in UKBB. Associations with fatty acid metabolites, which were also observed for LC incidence, may reflect dietary risk factors, or reflect metabolic shifts years before diagnosis, including increased β-oxidation (16, 63–65). We have previously reported inverse associations of lipid fractions in VLDL with BC incidence in premenopausal women (29), and this finding was replicated in premenopausal women in UKBB in the current study.

Positive associations of ketone bodies—acetoacetate, acetone, and β-hydroxybutyrate—were observed for all cancers in our study. Ketone bodies result mainly from fatty acid oxidation in the liver, and changes in ketone bodies may reflect metabolic reprogramming that both may promote or protect against cancer (66, 67).

Chronic low-grade inflammation is a well-known risk factor for cancer development, and tumor-promoting inflammation is one of the hallmarks of cancer (18). Blood levels of GlycA products are emerging as biomarkers for systemic inflammation (68–70). Our analyses indicated that elevated GlycA levels were associated with increased incidences of both LC and CRC. Similarly, low blood concentration of albumin may reflect inflammation (71), and albumin was found to be inversely associated with incidence of LC, CRC and PCa. Surprisingly, neither GlycA nor albumin was associated with incidence of BC. Our results are not in agreement with a previous prospective case-cohort study in the EPIC cohort, which reported an inverse association between albumin levels and BC incidence, but not with PCa, LC, or CRC incidence (72). Thus, further studies are needed to determine the association between albumin and cancer risk.

Dietary factors may influence several circulating metabolite levels, including glycolysis metabolites, lipids, fatty acids, and glycerol-derived compounds which are all affected by dietary fat and carbohydrate intake (73–76). High-fat diets tend to elevate circulating lipid metabolites, whereas low-fat or calorie-restricted diets may reduce their availability. In this study, we did not account for dietary intake for two main reasons —a) due to the differences in the variables and questionnaires used to capture diet and lifestyle features between HUNT3 and UKBB and b) dietary questionnaires are predominantly self-reported, which may introduce a response bias. However, future investigations incorporating detailed dietary information are warranted.

As metabolite levels were measured in blood samples collected years before diagnosis in our study, the observed associations between metabolite concentrations and cancer incidence may reflect broader metabolic alterations that promote the development and growth of cancer cells. Comparing results from metabolomics studies poses certain challenges due to variations in population characteristics and analytical techniques (77). Metabolites in both cohorts were quantified by Nightingale Health; however, different blood sample types were used: EDTA-plasma in UKBB and serum in HUNT3, which may influence metabolite measurements. Nonetheless, replicating potential associations—both for individual metabolites and more broadly across metabolite classes—is crucial for reliable interpretation of findings. While metabolic associations in UKBB and HUNT3 were mainly showing the same trends for LC and CRC, striking discrepancies were observed for PCa, suggesting potential cohort-specific differences in population characteristics, data collection methods, or underlying biological mechanisms.

The key strengths of this study include: 1) the utilization of two distinct and large population-based biobanks, 2) the use of NMR metabolomics, a highly robust method for quantifying metabolites and lipoproteins, and 3) a prospective design with extended follow-up. Additionally, we conducted a sensitivity analysis by excluding participants with any form of cancer prior to or at recruitment. The associations remained consistent, indicating no confounding influence from other malignancies on the observed relationships. The metabolic profiles for both UKBB and HUNT3 were measured in a non-fasting state, with only a one-hour difference in the average time since last meal, which likely had minimal impact, maintaining cohort comparability. However, differences in dietary habits between UKBB and HUNT3 could still pose a comparability issue. Previous studies have demonstrated that lipid levels exhibit minimal changes in response to typical food intake in the general population (73–76, 78), and that non-fasting lipid profiles could reliably predict an increased risk of cardiovascular events (79). However, some small-molecular metabolites are very sensitive to diet and may be affected by the time since last meal (80). Although HUNT3 mirrored the associations observed in UKBB for LC incidence, the lower number of cases in HUNT3 may have contributed to less precise results. The incidence of PCa in HUNT3 (5.9%) was higher than in UKBB (4.6%). This difference may explain the discrepancy in the direction of associations in the two cohorts, potentially due to varying baseline risk profiles or differing environmental and genetic factors influencing disease risk. Moreover, UKBB represents an older population, while HUNT3 encompasses a broader age range (18–100 years vs. 40–69 years). Additionally, the low participation rate in UKBB (5.5%) may have introduced selection bias, as participants tend to be healthier than non-participants (81). In contrast, HUNT3, with a higher participation rate (54.1%), is considered more representative of the general population (82).

## Conclusion

This prospective metabolomic study examined the associations between blood metabolites and the incidence of breast, prostate, lung and colorectal cancers in two large population biobanks. The study highlights inverse associations between most lipids in LDL and IDL subfractions and lung cancer incidence, predominantly observed in the UK Biobank. Similar associations were observed for albumin and some fatty acids. Ketone bodies, including acetoacetate, acetone, and β-hydroxybutyrate, as well as the inflammation-related metabolite glycoprotein acetylation, showed positive associations with incidence of lung cancer. Although the direction of these associations was generally consistent in the HUNT Study, none reached statistical significance, likely due to the smaller sample size.

### Supplementary material

Supplementary data is available on GitHub.

### Ethics declarations

The UKBB study was approved by the Northwest Multi-Centre Research Ethics Committee (reference number 11/NW/0382). The use of UKBB data in the current study was approved by the Regional Committees for Medical and Health Research Ethics (REK 427916). All participants signed an informed consent form. The UKBB database was made in accordance with the Declaration of Helsinki. The HUNT Study was approved by the Regional Committee for Ethics in Medical Research (REK; reference number 468380). All participants signed informed written consent on participation in HUNT, with linkage to previous HUNT surveys and specific registries in accordance with the Declaration of Helsinki.

## Acknowledgement

This research has been conducted using data from UK Biobank, a major biomedical database (https://www.ukbiobank.ac.uk/), under application number 40135. The Trøndelag Health Study (The HUNT Study) is a collaboration between HUNT Research Center (Faculty of Medicine and Health Sciences, NTNU, Norwegian University of Science and Technology), Trøndelag County Council, Central Norway Regional Health Authority, and the Norwegian Institute of Public Health.

## Author contributions

MD, AS, BMB, JD and GFG designed the study. MD and AS performed the statistical analysis and drafted the manuscript. This publication is the work of the authors. All authors read and approved the final manuscript.

## Data availability

The UK Biobank data are available to researchers, subject to successful registration and application process via their website (https://www.ukbiobank.ac.uk/). Data from the HUNT Study used in research projects will, when reasonably requested by others, be made available on request to the HUNT Data Access Committee. The HUNT data access information describes the policy regarding data availability (https://www.ntnu.edu/hunt/data).

