## Supplementary File for "Associations between circulating metabolites and incidence of breast, prostate, lung and colorectal cancers in the HUNT Study and UK Biobank"

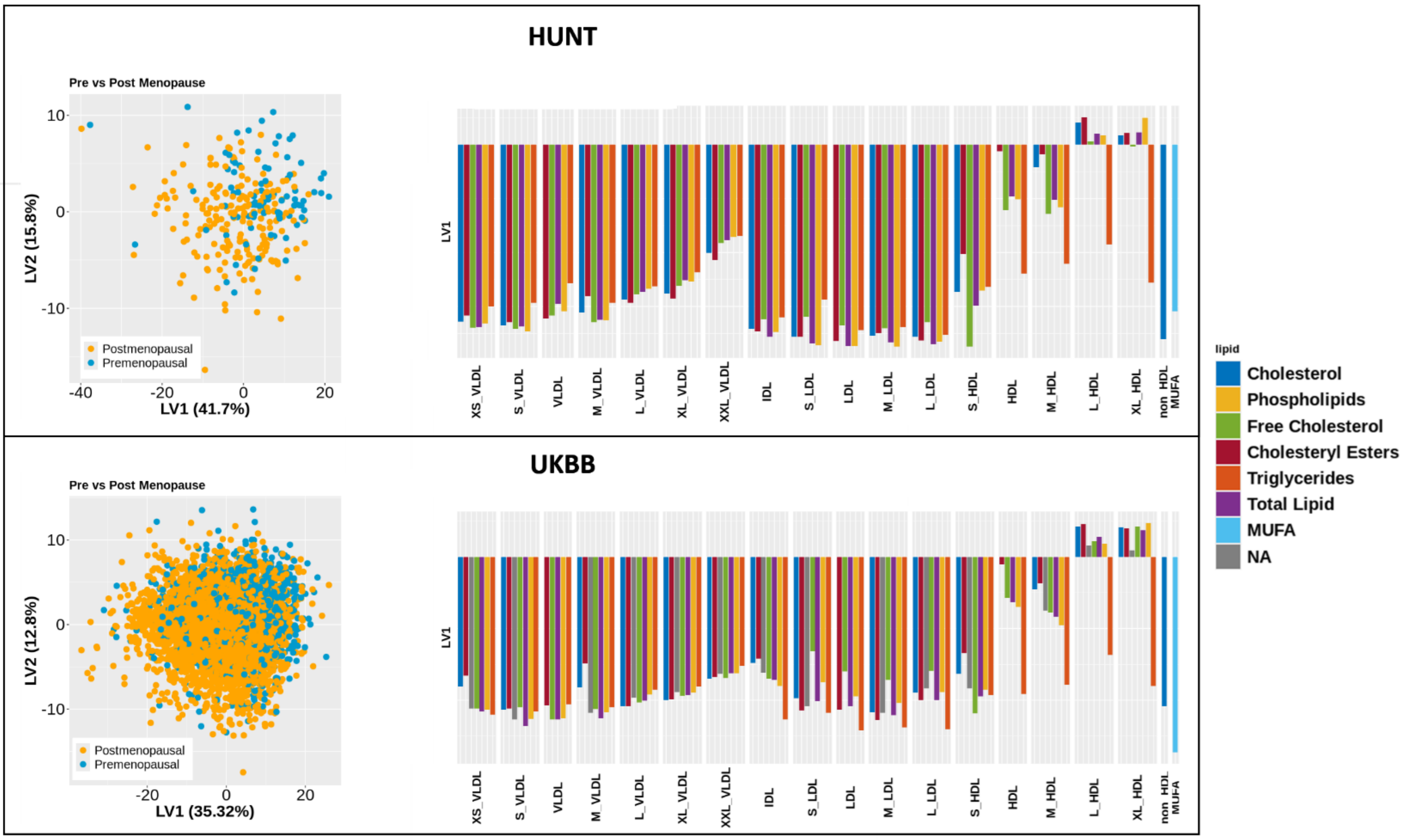


**Supplementary Figure S1**. PLS-DA analysis showed difference in the metabolic profiles of premenopausal (blue) and postmenopausal (yellow) future breast cancer patients in both HUNT3 (top left) and UKBB (bottom left). A clear separation is observed in the score plot by incorporating the first two latent variables LV1 and LV2. The direction of change in the profile remains consistent for both HUNT3 (top right) and UKBB (bottom right), where the elevated levels of most lipids subfraction was observed in postmenopausal women. The average area under the receiver operating curve obtained in 5-fold cross-validation are inscribed in the score plots.

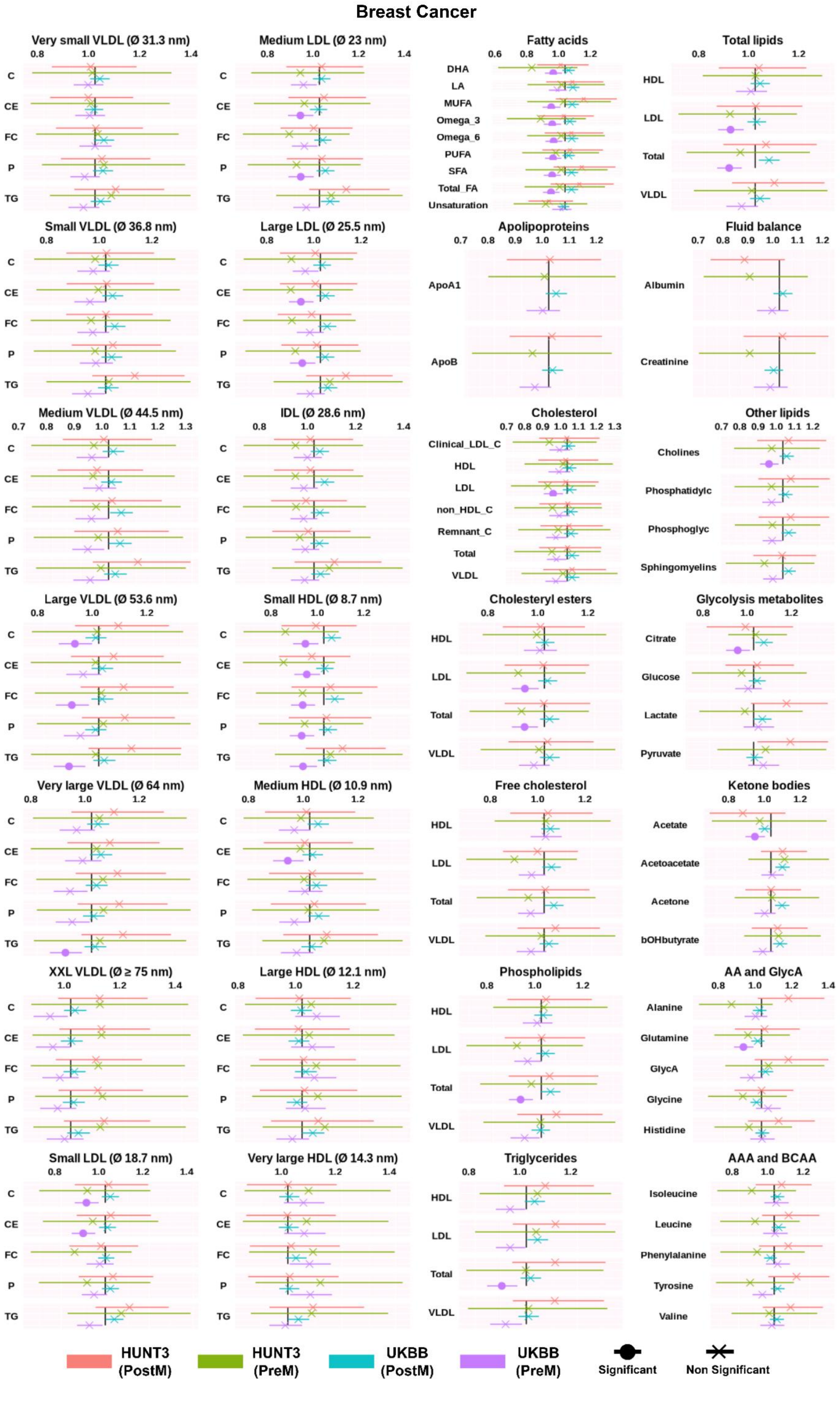

Breast Cancer
Very small VLDL (Ø 31.3 nm)
Medium LDL (Ø 23 nm)
Fatty acids
Total lipids
Small VLDL (Ø 36.8 nm)
Large LDL (Ø 25.5 nm)
Apolipoproteins
Fluid balance
Medium VLDL (Ø 44.5 nm)
IDL (Ø 28.6 nm)
Cholesterol
Other lipids
Large VLDL (Ø 53.6 nm)
Small HDL (Ø 8.7 nm)
Cholesteryl esters
Glycolysis metabolites
Very large VLDL (Ø 64 nm)
Medium HDL (Ø 10.9 nm)
Free cholesterol
Ketone bodies
XXL VLDL (Ø ≥ 75 nm)
Large HDL (Ø 12.1 nm)
Phospholipids
AA and GlycA
Small LDL (Ø 18.7 nm)
Very large HDL (Ø 14.3 nm)
Triglycerides
AAA and BCAA
C
CE
FC
P
TG
DHA
LA
MUFA
Omega_3
Omega_6
PUFA
SFA
Total_FA
Unsaturation
HDL
LDL
Total
VLDL
ApoA1
ApoB
Albumin
Creatinine
Clinical_LDL_C
non_HDL_C
Remnant_C
Cholines
Phosphatidylc
Phosphoglyc
Sphingomyelins
Citrate
Glucose
Lactate
Pyruvate
Acetate
Acetoacetate
Acetone
bOHbutyrate
Alanine
Glutamine
GlycA
Glycine
Histidine
Isoleucine
Leucine
Phenylalanine
Tyrosine
Valine
HUNT3 (PostM)
HUNT3 (PreM)
UKBB (PostM)
UKBB (PreM)
Significant
Non Significant

**Supplementary Figure S2**. Hazard ratios (HR) of BC incidence in relation to metabolites in menopause status based stratified cohort were assessed in HUNT3 (events = 86 and 197 in pre- and post-menopausal women respectively, n = 9,129) and UKBB (events = 1,090 and 3,426 in pre- and post-menopausal women respectively, n = 139,684). The data are presented as HRs per 1-SD increment with 95% confidence intervals (CIs), based on SD-scaled concentrations. Cox proportional hazards regression models were adjusted for UKBB assessment center (for UKBB), BMI, alcohol consumption, smoking status, menarche status, and number of live births, with age used as the timescale. AA=Amino acids, AAA = Aromatic amino acids, ApoA1 = Apolipoprotein A1, ApoB = Apolipoprotein B, Albumin, bOHbutyrate = Beta-hydroxybutyrate, BCAA = Branched-chain amino acids, CE = Cholesteryl esters, C = Cholesterol, Clinical_LDL_C, Creatinine, DHA = Docosahexaenoic acid, FC = Free cholesterol, GlycA = Glycoprotein Acetylation, HDL = High-density lipoprotein, LA = Linoleic acid, LDL = Low-density lipoprotein, non_HDL_C, MUFA = Monounsaturated fatty acids, Phosphatidylc = Phosphatidylcholine, Phosphoglyc = Phosphoglycerides, P = Phospholipids, PUFA = Polyunsaturated fatty acids, Remnant_C, SFA = Saturated fatty acids, TG = Triglycerides, Total FA = Total fatty acids, VLDL = Very low-density lipoprotein, XXL = Extremely large, Ø = Diameter.

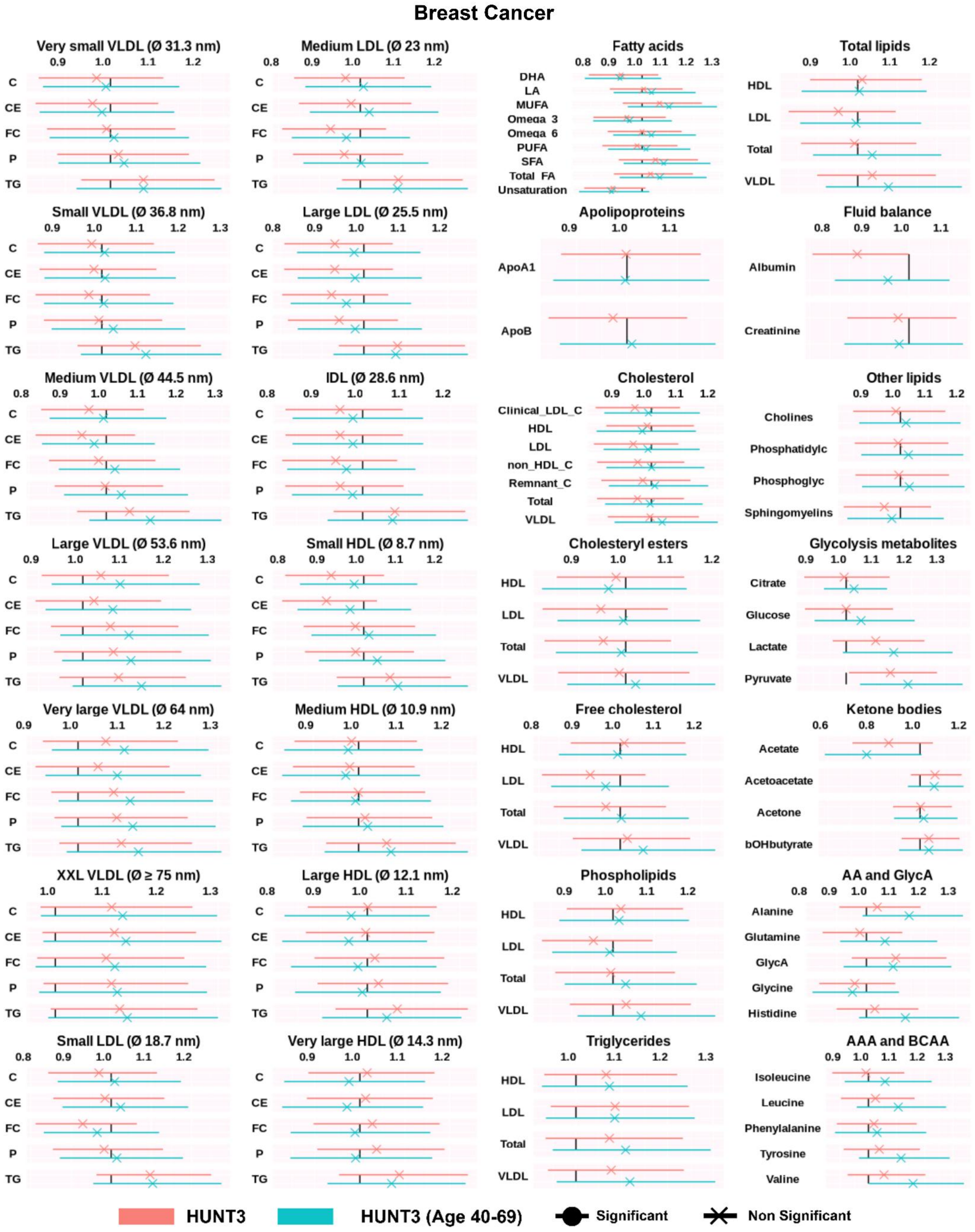
Breast Cancer
Very small VLDL (Ø 31.3 nm)
Medium LDL (Ø 23 nm)
Fatty acids
Total lipids
Small VLDL (Ø 36.8 nm)
Large LDL (Ø 25.5 nm)
Apolipoproteins
Fluid balance
Medium VLDL (Ø 44.5 nm)
IDL (Ø 28.6 nm)
Cholesterol
Other lipids
Large VLDL (Ø 53.6 nm)
Small HDL (Ø 8.7 nm)
Cholesteryl esters
Glycolysis metabolites
Very large VLDL (Ø 64 nm)
Medium HDL (Ø 10.9 nm)
Free cholesterol
Ketone bodies
XXL VLDL (Ø ≥ 75 nm)
Large HDL (Ø 12.1 nm)
Phospholipids
AA and GlycA
Small LDL (Ø 18.7 nm)
Very large HDL (Ø 14.3 nm)
Triglycerides
AAA and BCAA
C
CE
FC
P
TG
DHA
LA
MUFA
Omega 3
Omega 6
PUFA
SFA
Total FA
Unsaturation
HDL
LDL
Total
VLDL
ApoA1
ApoB
Albumin
Creatinine
Clinical_LDL_C
non_HDL_C
Remnant_C
Cholines
Phosphatidylc
Phosphoglyc
Sphingomyelins
Citrate
Glucose
Lactate
Pyruvate
Acetate
Acetoacetate
Acetone
bOHbutyrate
Alanine
Glutamine
GlycA
Glycine
Histidine
Isoleucine
Leucine
Phenylalanine
Tyrosine
Valine
HUNT3
HUNT3 (Age 40-69)
Significant
Non Significant

**Supplementary Figure S3**. Comparing the hazard ratios (HR) of breast cancer incidence in relation to metabolites between HUNT3 and age-restricted HUNT3. The data are presented as HRs per 1-SD increment with 95% confidence intervals (CIs), based on SD-scaled concentrations. Cox proportional hazards regression models were adjusted for BMI, alcohol consumption, smoking status, menarche status, and number of live births, with age used as the timescale. AA=Amino acids, AAA = Aromatic amino acids, ApoA1 = Apolipoprotein A1, ApoB = Apolipoprotein B, Albumin, bOHbutyrate = Beta-hydroxybutyrate, BCAA = Branched-chain amino acids, CE = Cholesteryl esters, C = Cholesterol, Clinical_LDL_C, Creatinine, DHA = Docosahexaenoic acid, FC = Free cholesterol, GlycA = Glycoprotein Acetylation, HDL = High-density lipoprotein, LA = Linoleic acid, LDL = Low-density lipoprotein, non_HDL_C, MUFA = Monounsaturated fatty acids, Phosphatidylc = Phosphatidylcholine, Phosphoglyc = Phosphoglycerides, P = Phospholipids, PUFA = Polyunsaturated fatty acids, Remnant_C, SFA = Saturated fatty acids, TG = Triglycerides, Total FA = Total fatty acids, VLDL = Very low-density lipoprotein, XXL = Extremely large, Ø = Diameter.

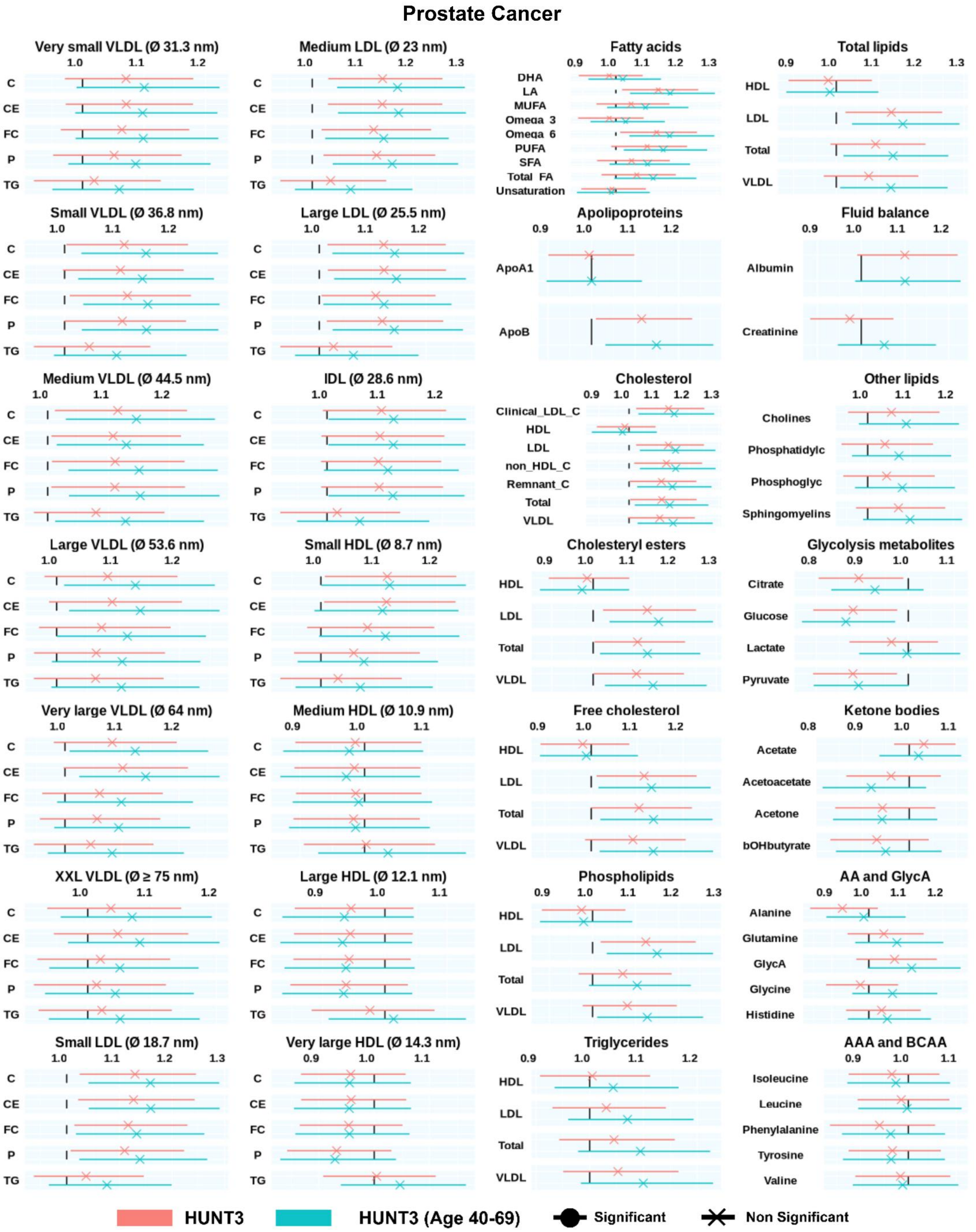

Prostate Cancer
Very small VLDL (Ø 31.3 nm)
1.0 1.1 1.2
C CE FC P TG
Medium LDL (Ø 23 nm)
1.0 1.1 1.2 1.3
C CE FC P TG
Fatty acids
0.9 1.0 1.1 1.2 1.3
DHA LA MUFA Omega_3 Omega_6 PUFA SFA Total_FA Unsaturation
Total lipids
0.9 1.0 1.1 1.2 1.3
HDL LDL Total VLDL
Small VLDL (Ø 36.8 nm)
1.0 1.1 1.2
C CE FC P TG
Large LDL (Ø 25.5 nm)
1.0 1.1 1.2
C CE FC P TG
Apolipoproteins
0.9 1.0 1.1 1.2
ApoA1 ApoB
Fluid balance
0.9 1.0 1.1 1.2
Albumin Creatinine
Medium VLDL (Ø 44.5 nm)
1.0 1.1 1.2
C CE FC P TG
IDL (Ø 28.6 nm)
1.0 1.1 1.2
C CE FC P TG
Cholesterol
0.9 1.0 1.1 1.2 1.3
Clinical_LDL_C HDL LDL non_HDL_C Remnant_C Total VLDL
Other lipids
1.0 1.1 1.2
Cholines Phosphatidylc Phosphoglyc Sphingomyelins
Large VLDL (Ø 53.6 nm)
1.0 1.1 1.2
C CE FC P TG
Small HDL (Ø 8.7 nm)
1.0 1.1 1.2
C CE FC P TG
Cholesteryl esters
0.9 1.0 1.1 1.2 1.3
HDL LDL Total VLDL
Glycolysis metabolites
0.8 0.9 1.0 1.1
Citrate Glucose Lactate Pyruvate
Very large VLDL (Ø 64 nm)
1.0 1.1 1.2
C CE FC P TG
Medium HDL (Ø 10.9 nm)
0.9 1.0 1.1
C CE FC P TG
Free cholesterol
0.9 1.0 1.1 1.2
HDL LDL Total VLDL
Ketone bodies
0.8 0.9 1.0 1.1
Acetate Acetoacetate Acetone bOHbutyrate
XXL VLDL (Ø ≥ 75 nm)
1.0 1.1 1.2
C CE FC P TG
Large HDL (Ø 12.1 nm)
0.9 1.0 1.1
C CE FC P TG
Phospholipids
0.9 1.0 1.1 1.2 1.3
HDL LDL Total VLDL
AA and GlycA
0.9 1.0 1.1 1.2
Alanine Glutamine GlycA Glycine Histidine
Small LDL (Ø 18.7 nm)
1.0 1.1 1.2 1.3
C CE FC P TG
Very large HDL (Ø 14.3 nm)
0.9 1.0 1.1
C CE FC P TG
Triglycerides
0.9 1.0 1.1 1.2
HDL LDL Total VLDL
AAA and BCAA
0.9 1.0 1.1
Isoleucine Leucine Phenylalanine Tyrosine Valine
HUNT3
HUNT3 (Age 40-69)
Significant
Non Significant

**Supplementary Figure S4**. Comparing the hazard ratios (HR) of prostate cancer incidence in relation to metabolites between HUNT3 and age-restricted HUNT3. The data are presented as HRs per 1-SD increment with 95% confidence intervals (CIs), based on SD-scaled concentrations. Cox proportional hazards regression models were adjusted for BMI, alcohol consumption, smoking status, marital status, with age used as the timescale. AA=Amino acids, AAA = Aromatic amino acids, ApoA1 = Apolipoprotein A1, ApoB = Apolipoprotein B, Albumin, bOHbutyrate = Beta-hydroxybutyrate, BCAA = Branched-chain amino acids, CE = Cholesteryl esters, C = Cholesterol, Clinical_LDL_C, Creatinine, DHA = Docosahexaenoic acid, FC = Free cholesterol, GlycA = Glycoprotein Acetylation, HDL = High-density lipoprotein, LA = Linoleic acid, LDL = Low-density lipoprotein, non_HDL_C, MUFA = Monounsaturated fatty acids, Phosphatidylc = Phosphatidylcholine, Phosphoglyc = Phosphoglycerides, P = Phospholipids, PUFA = Polyunsaturated fatty acids, Remnant_C, SFA = Saturated fatty acids, TG = Triglycerides, Total FA = Total fatty acids, VLDL = Very low-density lipoprotein, XXL = Extremely large, Ø = Diameter.

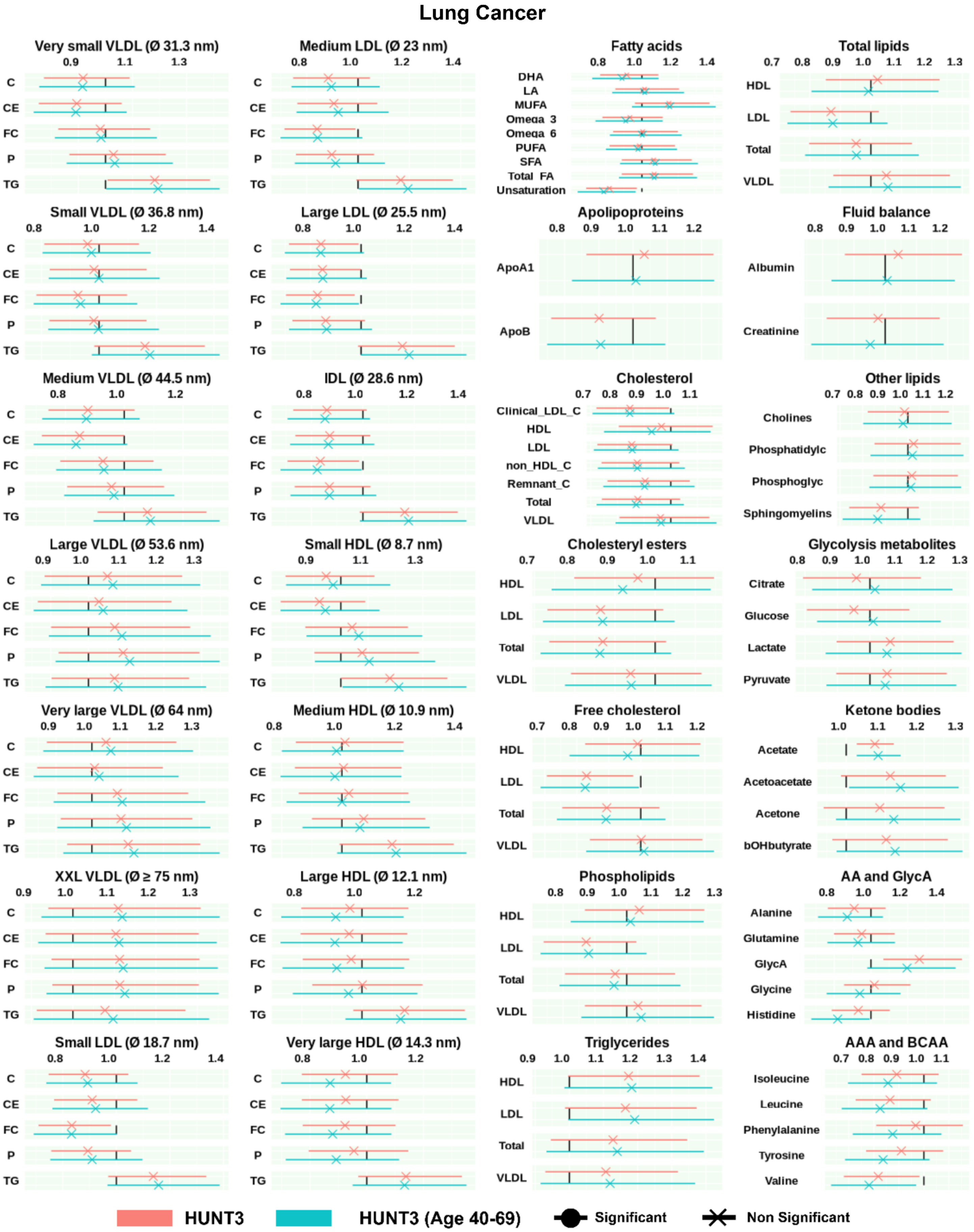

Lung Cancer
Very small VLDL (Ø 31.3 nm)
Medium LDL (Ø 23 nm)
Fatty acids
Total lipids
Small VLDL (Ø 36.8 nm)
Large LDL (Ø 25.5 nm)
Apolipoproteins
Fluid balance
Medium VLDL (Ø 44.5 nm)
IDL (Ø 28.6 nm)
Cholesterol
Other lipids
Large VLDL (Ø 53.6 nm)
Small HDL (Ø 8.7 nm)
Cholesteryl esters
Glycolysis metabolites
Very large VLDL (Ø 64 nm)
Medium HDL (Ø 10.9 nm)
Free cholesterol
Ketone bodies
XXL VLDL (Ø ≥ 75 nm)
Large HDL (Ø 12.1 nm)
Phospholipids
AA and GlycA
Small LDL (Ø 18.7 nm)
Very large HDL (Ø 14.3 nm)
Triglycerides
AAA and BCAA
C
CE
FC
P
TG
DHA
LA
MUFA
Omega_3
Omega_6
PUFA
SFA
Total_FA
Unsaturation
HDL
LDL
Total
VLDL
ApoA1
ApoB
Albumin
Creatinine
Clinical_LDL_C
non_HDL_C
Remnant_C
Cholines
Phosphatidylc
Phosphoglyc
Sphingomyelins
Citrate
Glucose
Lactate
Pyruvate
Acetate
Acetoacetate
Acetone
bOHbutyrate
Alanine
Glutamine
GlycA
Glycine
Histidine
Isoleucine
Leucine
Phenylalanine
Tyrosine
Valine
HUNT3
HUNT3 (Age 40-69)
Significant
Non Significant

**Supplementary Figure S5**. Comparing the hazard ratios (HR) of lung cancer incidence in relation to metabolites between HUNT3 and age-restricted HUNT3. The data are presented as HRs per 1-SD increment with 95% confidence intervals (CIs), based on SD-scaled concentrations. Cox proportional hazards regression models were adjusted for sex, BMI, alcohol consumption, smoking status, marital status, with age used as the timescale. AA=Amino acids, AAA = Aromatic amino acids, ApoA1 = Apolipoprotein A1, ApoB = Apolipoprotein B, Albumin, bOHbutyrate = Beta-hydroxybutyrate, BCAA = Branched-chain amino acids, CE = Cholesteryl esters, C = Cholesterol, Clinical_LDL_C, Creatinine, DHA = Docosahexaenoic acid, FC = Free cholesterol, GlycA = Glycoprotein Acetylation, HDL = High-density lipoprotein, LA = Linoleic acid, LDL = Low-density lipoprotein, non_HDL_C, MUFA = Monounsaturated fatty acids, Phosphatidylc = Phosphatidylcholine, Phosphoglyc = Phosphoglycerides, P = Phospholipids, PUFA = Polyunsaturated fatty acids, Remnant_C, SFA = Saturated fatty acids, TG = Triglycerides, Total FA = Total fatty acids, VLDL = Very low-density lipoprotein, XXL = Extremely large, Ø = Diameter.

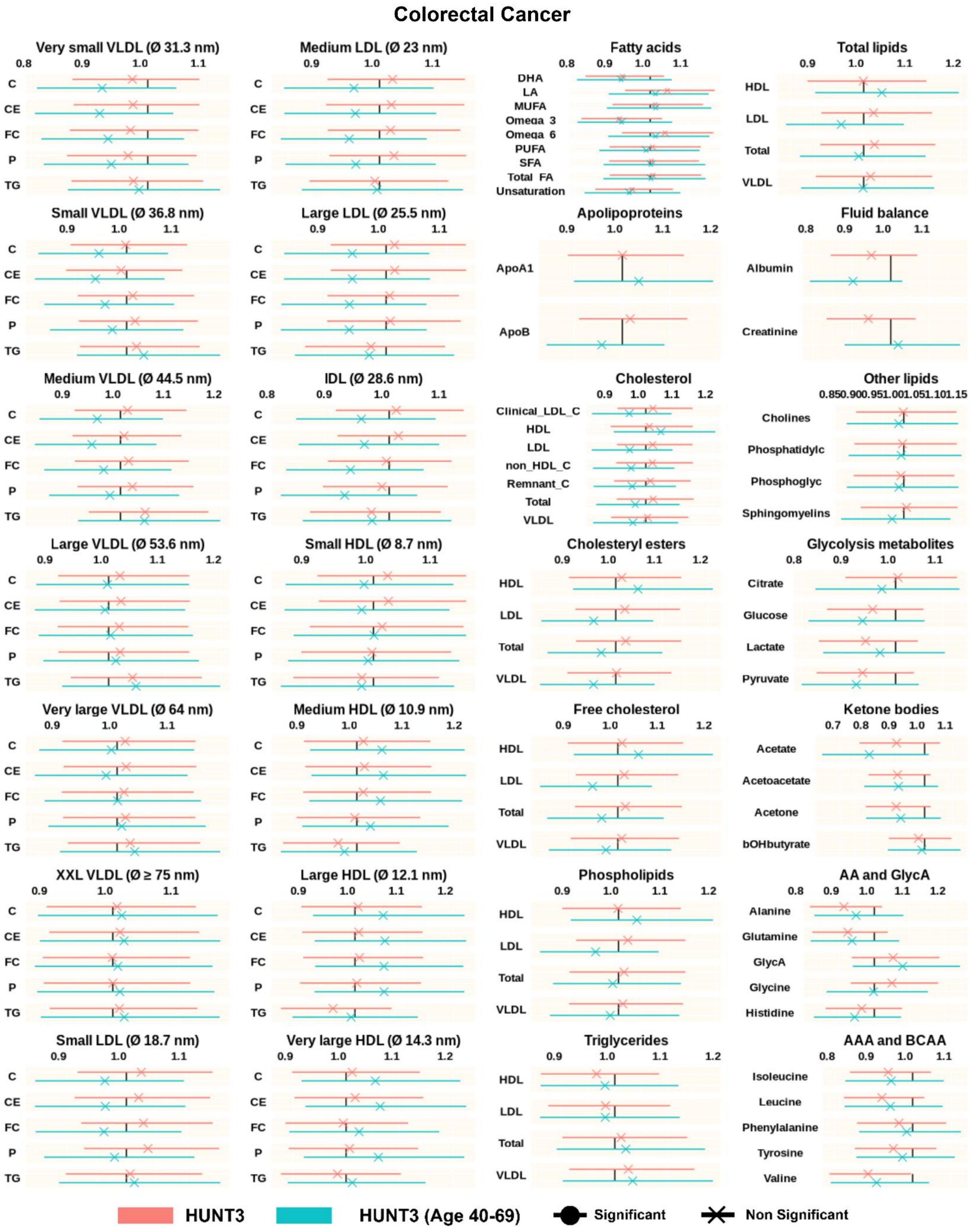

Colorectal Cancer
Very small VLDL (Ø 31.3 nm)
Medium LDL (Ø 23 nm)
Fatty acids
Total lipids
Small VLDL (Ø 36.8 nm)
Large LDL (Ø 25.5 nm)
Apolipoproteins
Fluid balance
Medium VLDL (Ø 44.5 nm)
IDL (Ø 28.6 nm)
Cholesterol
Other lipids
Large VLDL (Ø 53.6 nm)
Small HDL (Ø 8.7 nm)
Cholesteryl esters
Glycolysis metabolites
Very large VLDL (Ø 64 nm)
Medium HDL (Ø 10.9 nm)
Free cholesterol
Ketone bodies
XXL VLDL (Ø ≥ 75 nm)
Large HDL (Ø 12.1 nm)
Phospholipids
AA and GlycA
Small LDL (Ø 18.7 nm)
Very large HDL (Ø 14.3 nm)
Triglycerides
AAA and BCAA
C
CE
FC
P
TG
DHA
LA
MUFA
Omega 3
Omega 6
PUFA
SFA
Total FA
Unsaturation
HDL
LDL
Total
VLDL
ApoA1
ApoB
Albumin
Creatinine
Clinical_LDL_C
non_HDL_C
Remnant_C
Cholines
Phosphatidylc
Phosphoglyc
Sphingomyelins
Citrate
Glucose
Lactate
Pyruvate
Acetate
Acetoacetate
Acetone
bOHbutyrate
Alanine
Glutamine
GlycA
Glycine
Histidine
Isoleucine
Leucine
Phenylalanine
Tyrosine
Valine
HUNT3
HUNT3 (Age 40-69)
Significant
Non Significant

**Supplementary Figure S6**. Comparing the hazard ratios (HR) of colorectal cancer incidence in relation to metabolites between HUNT3 and age-restricted HUNT3. The data are presented as HRs per 1-SD increment with 95% confidence intervals (CIs), based on SD-scaled concentrations. Cox proportional hazards regression models were adjusted for sex, BMI, alcohol consumption, smoking status, marital status, with age used as the timescale. AA=Amino acids, AAA = Aromatic amino acids, ApoA1 = Apolipoprotein A1, ApoB = Apolipoprotein B, Albumin, bOHbutyrate = Beta-hydroxybutyrate, BCAA = Branched-chain amino acids, CE = Cholesteryl esters, C = Cholesterol, Clinical_LDL_C, Creatinine, DHA = Docosahexaenoic acid, FC = Free cholesterol, GlycA = Glycoprotein Acetylation, HDL = High-density lipoprotein, LA = Linoleic acid, LDL = Low-density lipoprotein, non_HDL_C, MUFA = Monounsaturated fatty acids, Phosphatidylc = Phosphatidylcholine, Phosphoglyc = Phosphoglycerides, P = Phospholipids, PUFA = Polyunsaturated fatty acids, Remnant_C, SFA = Saturated fatty acids, TG = Triglycerides, Total FA = Total fatty acids, VLDL = Very low-density lipoprotein, XXL = Extremely large, Ø = Diameter.

**Supplementary Table S1.** Metabolites and their corresponding QCs included in the study from UKBB.

| Metabolites | | Corresponding Quality Control Variable | |
|---|---|---|---|
| **FID** | **Description** | **FID** | **Description** |
| 23474 | 3-Hydroxybutyrate | 23774 | 3-Hydroxybutyrate, QC Flag |
| 23475 | Acetate | 23775 | Acetate, QC Flag |
| 23476 | Acetoacetate | 23776 | Acetoacetate, QC Flag |
| 23477 | Acetone | 23777 | Acetone, QC Flag |
| 23460 | Alanine | 23760 | Alanine, QC Flag |
| 23479 | Albumin | 23779 | Albumin, QC Flag |
| 23440 | Apolipoprotein A1 | 23740 | Apolipoprotein A1, QC Flag |
| 23439 | Apolipoprotein B | 23739 | Apolipoprotein B, QC Flag |
| 23484 | Cholesterol in Chylomicrons and Extremely Large VLDL | 23784 | Cholesterol in Chylomicrons and Extremely Large VLDL, QC Flag |
| 23526 | Cholesterol in IDL | 23826 | Cholesterol in IDL, QC Flag |
| 23561 | Cholesterol in Large HDL | 23861 | Cholesterol in Large HDL, QC Flag |
| 23533 | Cholesterol in Large LDL | 23833 | Cholesterol in Large LDL, QC Flag |
| 23498 | Cholesterol in Large VLDL | 23798 | Cholesterol in Large VLDL, QC Flag |
| 23568 | Cholesterol in Medium HDL | 23868 | Cholesterol in Medium HDL, QC Flag |
| 23540 | Cholesterol in Medium LDL | 23840 | Cholesterol in Medium LDL, QC Flag |
| 23505 | Cholesterol in Medium VLDL | 23805 | Cholesterol in Medium VLDL, QC Flag |
| 23575 | Cholesterol in Small HDL | 23875 | Cholesterol in Small HDL, QC Flag |
| 23547 | Cholesterol in Small LDL | 23847 | Cholesterol in Small LDL, QC Flag |
| 23512 | Cholesterol in Small VLDL | 23812 | Cholesterol in Small VLDL, QC Flag |
| 23554 | Cholesterol in Very Large HDL | 23854 | Cholesterol in Very Large HDL, QC Flag |
| 23491 | Cholesterol in Very Large VLDL | 23791 | Cholesterol in Very Large VLDL, QC Flag |
| 23519 | Cholesterol in Very Small VLDL | 23819 | Cholesterol in Very Small VLDL, QC Flag |
| 23485 | Cholesteryl Esters in Chylomicrons and Extremely Large VLDL | 23785 | Cholesteryl Esters in Chylomicrons and Extremely Large VLDL, QC Flag |
| 23418 | Cholesteryl Esters in HDL | 23718 | Cholesteryl Esters in HDL, QC Flag |
| 23527 | Cholesteryl Esters in IDL | 23827 | Cholesteryl Esters in IDL, QC Flag |
| 23417 | Cholesteryl Esters in LDL | 23717 | Cholesteryl Esters in LDL, QC Flag |
| 23562 | Cholesteryl Esters in Large HDL | 23862 | Cholesteryl Esters in Large HDL, QC Flag |
| 23534 | Cholesteryl Esters in Large LDL | 23834 | Cholesteryl Esters in Large LDL, QC Flag |
| 23499 | Cholesteryl Esters in Large VLDL | 23799 | Cholesteryl Esters in Large VLDL, QC Flag |
| 23569 | Cholesteryl Esters in Medium HDL | 23869 | Cholesteryl Esters in Medium HDL, QC Flag |
| 23541 | Cholesteryl Esters in Medium LDL | 23841 | Cholesteryl Esters in Medium LDL, QC Flag |
| 23506 | Cholesteryl Esters in Medium VLDL | 23806 | Cholesteryl Esters in Medium VLDL, QC Flag |
| 23576 | Cholesteryl Esters in Small HDL | 23876 | Cholesteryl Esters in Small HDL, QC Flag |

| | | | |
|---|---|---|---|
| 23548 | Cholesteryl Esters in Small LDL | 23848 | Cholesteryl Esters in Small LDL, QC Flag |
| 23513 | Cholesteryl Esters in Small VLDL | 23813 | Cholesteryl Esters in Small VLDL, QC Flag |
| 23416 | Cholesteryl Esters in VLDL | 23716 | Cholesteryl Esters in VLDL, QC Flag |
| 23555 | Cholesteryl Esters in Very Large HDL | 23855 | Cholesteryl Esters in Very Large HDL, QC Flag |
| 23492 | Cholesteryl Esters in Very Large VLDL | 23792 | Cholesteryl Esters in Very Large VLDL, QC Flag |
| 23520 | Cholesteryl Esters in Very Small VLDL | 23820 | Cholesteryl Esters in Very Small VLDL, QC Flag |
| 23473 | Citrate | 23773 | Citrate, QC Flag |
| 23404 | Clinical LDL Cholesterol | 23704 | Clinical LDL Cholesterol, QC Flag |
| 23478 | Creatinine | 23778 | Creatinine, QC Flag |
| 23443 | Degree of Unsaturation | 23743 | Degree of Unsaturation, QC Flag |
| 23450 | Docosahexaenoic Acid | 23750 | Docosahexaenoic Acid, QC Flag |
| 23486 | Free Cholesterol in Chylomicrons and Extremely Large VLDL | 23786 | Free Cholesterol in Chylomicrons and Extremely Large VLDL, QC Flag |
| 23422 | Free Cholesterol in HDL | 23722 | Free Cholesterol in HDL, QC Flag |
| 23528 | Free Cholesterol in IDL | 23828 | Free Cholesterol in IDL, QC Flag |
| 23421 | Free Cholesterol in LDL | 23721 | Free Cholesterol in LDL, QC Flag |
| 23563 | Free Cholesterol in Large HDL | 23863 | Free Cholesterol in Large HDL, QC Flag |
| 23535 | Free Cholesterol in Large LDL | 23835 | Free Cholesterol in Large LDL, QC Flag |
| 23500 | Free Cholesterol in Large VLDL | 23800 | Free Cholesterol in Large VLDL, QC Flag |
| 23570 | Free Cholesterol in Medium HDL | 23870 | Free Cholesterol in Medium HDL, QC Flag |
| 23542 | Free Cholesterol in Medium LDL | 23842 | Free Cholesterol in Medium LDL, QC Flag |
| 23507 | Free Cholesterol in Medium VLDL | 23807 | Free Cholesterol in Medium VLDL, QC Flag |
| 23577 | Free Cholesterol in Small HDL | 23877 | Free Cholesterol in Small HDL, QC Flag |
| 23549 | Free Cholesterol in Small LDL | 23849 | Free Cholesterol in Small LDL, QC Flag |
| 23514 | Free Cholesterol in Small VLDL | 23814 | Free Cholesterol in Small VLDL, QC Flag |
| 23420 | Free Cholesterol in VLDL | 23720 | Free Cholesterol in VLDL, QC Flag |
| 23556 | Free Cholesterol in Very Large HDL | 23856 | Free Cholesterol in Very Large HDL, QC Flag |
| 23493 | Free Cholesterol in Very Large VLDL | 23793 | Free Cholesterol in Very Large VLDL, QC Flag |
| 23521 | Free Cholesterol in Very Small VLDL | 23821 | Free Cholesterol in Very Small VLDL, QC Flag |
| 23470 | Glucose | 23770 | Glucose, QC Flag |
| 23461 | Glutamine | 23761 | Glutamine, QC Flag |
| 23462 | Glycine | 23762 | Glycine, QC Flag |
| 23480 | Glycoprotein Acetyls | 23780 | Glycoprotein Acetyls, QC Flag |
| 23406 | HDL Cholesterol | 23706 | HDL Cholesterol, QC Flag |
| 23463 | Histidine | 23763 | Histidine, QC Flag |
| 23465 | Isoleucine | 23765 | Isoleucine, QC Flag |

| | | | |
|---|---|---|---|
| 23405 | LDL Cholesterol | 23705 | LDL Cholesterol, QC Flag |
| 23471 | Lactate | 23771 | Lactate, QC Flag |
| 23466 | Leucine | 23766 | Leucine, QC Flag |
| 23449 | Linoleic Acid | 23749 | Linoleic Acid, QC Flag |
| 23447 | Monounsaturated Fatty Acids | 23747 | Monounsaturated Fatty Acids, QC Flag |
| 23444 | Omega-3 Fatty Acids | 23744 | Omega-3 Fatty Acids, QC Flag |
| 23445 | Omega-6 Fatty Acids | 23745 | Omega-6 Fatty Acids, QC Flag |
| 23468 | Phenylalanine | 23768 | Phenylalanine, QC Flag |
| 23437 | Phosphatidylcholines | 23737 | Phosphatidylcholines, QC Flag |
| 23434 | Phosphoglycerides | 23734 | Phosphoglycerides, QC Flag |
| 23483 | Phospholipids in Chylomicrons and Extremely Large VLDL | 23783 | Phospholipids in Chylomicrons and Extremely Large VLDL, QC Flag |
| 23414 | Phospholipids in HDL | 23714 | Phospholipids in HDL, QC Flag |
| 23525 | Phospholipids in IDL | 23825 | Phospholipids in IDL, QC Flag |
| 23413 | Phospholipids in LDL | 23713 | Phospholipids in LDL, QC Flag |
| 23560 | Phospholipids in Large HDL | 23860 | Phospholipids in Large HDL, QC Flag |
| 23532 | Phospholipids in Large LDL | 23832 | Phospholipids in Large LDL, QC Flag |
| 23497 | Phospholipids in Large VLDL | 23797 | Phospholipids in Large VLDL, QC Flag |
| 23567 | Phospholipids in Medium HDL | 23867 | Phospholipids in Medium HDL, QC Flag |
| 23539 | Phospholipids in Medium LDL | 23839 | Phospholipids in Medium LDL, QC Flag |
| 23504 | Phospholipids in Medium VLDL | 23804 | Phospholipids in Medium VLDL, QC Flag |
| 23574 | Phospholipids in Small HDL | 23874 | Phospholipids in Small HDL, QC Flag |
| 23546 | Phospholipids in Small LDL | 23846 | Phospholipids in Small LDL, QC Flag |
| 23511 | Phospholipids in Small VLDL | 23811 | Phospholipids in Small VLDL, QC Flag |
| 23412 | Phospholipids in VLDL | 23712 | Phospholipids in VLDL, QC Flag |
| 23553 | Phospholipids in Very Large HDL | 23853 | Phospholipids in Very Large HDL, QC Flag |
| 23490 | Phospholipids in Very Large VLDL | 23790 | Phospholipids in Very Large VLDL, QC Flag |
| 23518 | Phospholipids in Very Small VLDL | 23818 | Phospholipids in Very Small VLDL, QC Flag |
| 23446 | Polyunsaturated Fatty Acids | 23746 | Polyunsaturated Fatty Acids, QC Flag |
| 23472 | Pyruvate | 23772 | Pyruvate, QC Flag |
| 23402 | Remnant Cholesterol (Non-HDL, Non-LDL -Cholesterol) | 23702 | Remnant Cholesterol (Non-HDL, Non-LDL -Cholesterol), QC Flag |
| 23448 | Saturated Fatty Acids | 23748 | Saturated Fatty Acids, QC Flag |
| 23438 | Sphingomyelins | 23738 | Sphingomyelins, QC Flag |
| 23400 | Total Cholesterol | 23701 | Total Cholesterol Minus HDL-C, QC Flag |
| 23401 | Total Cholesterol Minus HDL-C | 23700 | Total Cholesterol, QC Flag |
| 23436 | Total Cholines | 23736 | Total Cholines, QC Flag |
| 23415 | Total Esterified Cholesterol | 23715 | Total Esterified Cholesterol, QC Flag |
| 23442 | Total Fatty Acids | 23742 | Total Fatty Acids, QC Flag |

| | | | |
|---|---|---|---|
| 23419 | Total Free Cholesterol | 23719 | Total Free Cholesterol, QC Flag |
| 23426 | Total Lipids in HDL | 23726 | Total Lipids in HDL, QC Flag |
| 23425 | Total Lipids in LDL | 23725 | Total Lipids in LDL, QC Flag |
| 23423 | Total Lipids in Lipoprotein Particles | 23723 | Total Lipids in Lipoprotein Particles, QC Flag |
| 23424 | Total Lipids in VLDL | 23724 | Total Lipids in VLDL, QC Flag |
| 23411 | Total Phospholipids in Lipoprotein Particles | 23711 | Total Phospholipids in Lipoprotein Particles, QC Flag |
| 23407 | Total Triglycerides | 23707 | Total Triglycerides, QC Flag |
| 23487 | Triglycerides in Chylomicrons and Extremely Large VLDL | 23787 | Triglycerides in Chylomicrons and Extremely Large VLDL, QC Flag |
| 23410 | Triglycerides in HDL | 23710 | Triglycerides in HDL, QC Flag |
| 23529 | Triglycerides in IDL | 23829 | Triglycerides in IDL, QC Flag |
| 23409 | Triglycerides in LDL | 23709 | Triglycerides in LDL, QC Flag |
| 23564 | Triglycerides in Large HDL | 23864 | Triglycerides in Large HDL, QC Flag |
| 23536 | Triglycerides in Large LDL | 23836 | Triglycerides in Large LDL, QC Flag |
| 23501 | Triglycerides in Large VLDL | 23801 | Triglycerides in Large VLDL, QC Flag |
| 23571 | Triglycerides in Medium HDL | 23871 | Triglycerides in Medium HDL, QC Flag |
| 23543 | Triglycerides in Medium LDL | 23843 | Triglycerides in Medium LDL, QC Flag |
| 23508 | Triglycerides in Medium VLDL | 23808 | Triglycerides in Medium VLDL, QC Flag |
| 23578 | Triglycerides in Small HDL | 23878 | Triglycerides in Small HDL, QC Flag |
| 23550 | Triglycerides in Small LDL | 23850 | Triglycerides in Small LDL, QC Flag |
| 23515 | Triglycerides in Small VLDL | 23815 | Triglycerides in Small VLDL, QC Flag |
| 23408 | Triglycerides in VLDL | 23708 | Triglycerides in VLDL, QC Flag |
| 23557 | Triglycerides in Very Large HDL | 23857 | Triglycerides in Very Large HDL, QC Flag |
| 23494 | Triglycerides in Very Large VLDL | 23794 | Triglycerides in Very Large VLDL, QC Flag |
| 23522 | Triglycerides in Very Small VLDL | 23822 | Triglycerides in Very Small VLDL, QC Flag |
| 23469 | Tyrosine | 23769 | Tyrosine, QC Flag |
| 23403 | VLDL Cholesterol | 23703 | VLDL Cholesterol, QC Flag |
| 23467 | Valine | 23767 | Valine, QC Flag |

**Supplementary Table S2.** Variable IDs from UKBB and HUNT3.

| Description | UKBB FID | HUNT ID |
|---|---|---|
| Age at death | 40007 | † |
| Underlying (primary) cause of death ICD10 | 40001 | † |
| Contributory (secondary) causes of death ICD10 | 40002 | † |
| Sex | 31 | HUNT(Sex) |
| Month of birth | 52 | - |
| Year of birth | 34 | HUNT(BirthYear) |
| Age at recruitment | 21022 | PartAg.NT3BLQ1 |
| Type of cancer ICD10 (C50.x) | 40006 | ● |
| Type of cancer ICD9 (174.x) | 40013 | ● |
| Date of cancer diagnosis | 40005 | ● |
| Age at menarche | 2714 | MenarcAg.NT3BLI |
| Had menopause | 2724 | - |
| Age at menopause (last menstrual period) | 3581 | MenopAg.NT3BLI |
| Number of live births | 2734 | DelivN.NT3BLI |
| Diagnoses - ICD9 | 41271 | ● |
| Diagnoses - ICD10 | 41270 | ● |
| Smoking status | 20116 | SmoStat.NT3BLQ1 |
| Alcohol intake frequency. | 1558 | AlcTotUnitW.NT3BLQ1 |
| Body mass index (BMI) | 21001 | - |
| Height | - | Hei.NT3BLM |
| Weight | - | Wei.NT3BLM |
| (-) Unavailable or not used; (†) Information from The Norwegian Cause of Death Registry; (●) Information from the Cancer Registry of Norway | | |
